# Visual Accessibility through Open Shelving: Impacts on Cognitive Load, Motivation, Physical Activity, and User Perception in Older Adults with Mild Cognitive Impairment

**DOI:** 10.1101/2025.05.21.25328033

**Authors:** Ibrahim Bilau, BonWoo Koo, Esther Fu, Wendy Chau, Hyeokhyen Kwon, Eunhwa Yang

## Abstract

Older adults with Mild Cognitive Impairment (MCI) often experience difficulty locating items in kitchen cabinets, limiting their experiences in the kitchen and with meal preparation. This study examined how open shelving kitchen cabinets, relative to closed cabinets, affects cognitive load, motivation, physical activity, and user perception among older adults with MCI. Eleven older adults completed meal preparation tasks in both conditions while data on eye-tracking, wrist accelerometry, step count, task duration, and Intrinsic Motivation Inventory were collected. Participant profiles were developed using standardized z-scores, and post-task interviews captured qualitative perspectives. Results showed modest improvements in cognitive load, intrinsic motivation, and task duration, with physical activity significantly decreasing under open shelving, suggesting greater physical efficiency. Individual profiles revealed diverse patterns, including high performers, strugglers, motivated but physically strained, and balanced or average participants. Interview findings highlighted that emotional and aesthetic concerns, like dust and visual clutter, influenced attitudes toward open shelving regardless of objective functional benefits. These findings suggest that while open shelving may reduce cognitive and physical demands and enhance motivation and task efficiency, personalized and flexible design solutions are critical to support cognitive aging-in-place effectively.

## Introduction

Cognitive aging involves a gradual decline in functions, such as memory, attention, executive function, and visuospatial function, which can significantly impact daily living for older adults (Petersen et al., 1999; Derbie et al., 2022). Mild cognitive impairment (MCI), a transitional stage between normal aging and dementia, affects 10–20% of individuals aged 65 and older (Alzheimers.gov, 2024; Petersen et al., 1999), necessitating supportive environments to maintain independence. Approximately 75% of adults over the age of 50 prefer aging in place, living safely and independently in their homes (Binette, 2021; Thomas & Blanchard, 2009), which highlights the need for home designs that accommodate cognitive and physical challenges.

Among home environments, kitchen design is particularly critical for supporting functional independence among older adults with MCI, particularly for instrumental activities of daily living (IADLs) like meal preparation (Guo & Sapra, 2020). Individuals with MCI experience cognitive challenges, including reduced working memory capacity, slower processing speed, and difficulties in spatial navigation, all of which increase cognitive load during multi-step tasks like cooking (Wang et al., 2022; Lussier et al., 2019). These challenges manifest as difficulties in remembering/recalling recipe steps, locating ingredients, and operating appliances safely (Guralnik & Ferrucci, 2003; Liverman et al., 2015). Additionally, co-occurring physical limitations, such as restricted mobility and reduced grip strength, further complicate tasks by hindering access to items stored on high shelves and limiting effective utensil use (Allan et al., 2005; Nagi, 1976). Prior studies have shown that older adults with MCI often struggle to locate seasonings, spices, and utensils in kitchen cabinets (King, 2023; Machry et al., 2025). A sensor-based study further revealed that individuals with MCI spend more time searching through kitchen cabinets and refrigerators than their cognitively healthy counterparts (Lussier et al., 2019), highlighting the need for targeted kitchen design interventions.

Universal design principles advocate for intuitive and accessible spaces that enhance everyday cognition and reduce dependence (Centre for Excellence in Universal Design, 2015). Open shelving, for example, may improve visual accessibility, facilitate item recognition, and consequently reduce cognitive load, though its efficacy for individuals with MCI remains untested (King, 2023). While physical accommodations in home design have been extensively studied, empirical evidence regarding cognitive accommodations in kitchen environments, particularly concerning the impact of open shelving on visual accessibility versus the potential cognitive overload from visual clutter, remains sparse (Czaja et al., 2018). To address this gap in aging-in-place research, this study investigates how open shelving, compared to closed cabinets, affects cognitive load, intrinsic motivation, physical activity levels, step count, and task duration among older adults with MCI.

This study employs Cognitive Load Theory (CLT) and Self-Determination Theory (SDT) to investigate the impact of kitchen design on older adults with MCI. CLT posits that cognitive resources are finite and that environmental design can either alleviate or exacerbate cognitive load, influencing task performance (Sweller, 2010). SDT asserts that intrinsic motivation, which implies engagement in activities for their inherent satisfaction, relies on the fulfillment of the psychological needs for autonomy, competence, and relatedness (Ryan & Deci, 2000). Kitchen designs that reduce cognitive load and bolster intrinsic motivation can potentially improve task efficiency and well-being in older adults with MCI (Rodrigues et al., 2023).

Key constructs were operationalized as follows: Cognitive load was measured using multiple eye-tracking metrics, including fixation duration and rate, saccade length and velocity, pupil dilation, and blink rate and latency, which reflect cognitive processing demands (Zagermann et al., 2016). Increased cognitive load correlates with longer fixation duration, lower fixation rate, longer saccade length, higher saccade velocity, greater pupil dilation, lower blink rate, and higher blink latency (Holmqvist et al., 2011; Zagermann et al., 2016). These complementary measures were integrated to provide a multidimensional characterization of cognitive load, thereby strengthening the construct validity and reliability of our findings. Intrinsic motivation was operationalized using the Intrinsic Motivation Inventory (IMI), which assesses subscales such as Interest/Enjoyment, Competence, and Pressure/Tension, which correspond to the core psychological needs identified by SDT (Ryan, 1982; McAuley et al., 1989). Physical activity intensity and step count, another key outcome, were measured via acceleration magnitude (e.g., 0.5–1.5 m/s² for light activity) captured by wrist-worn sensors. This approach enabled precise quantification of physical effort during meal preparation tasks that vary with kitchen design features like open shelving compared to closed cabinets (Freedson et al., 1998).

This study addresses the following primary research questions:

1. Does open shelving reduce cognitive load in older adults with MCI?
2. Does open shelving enhance intrinsic motivation?
3. Does open shelving reduce physical effort during meal preparation, as measured by activity intensity and step count?
4. Does open shelving reduce task duration (improve task efficiency)?
5. Do individual participant profiles reflect variability in cognitive, motivational, and physical responses to open shelving?
6. Do the subjective perceptions of older adults with MCI differ from objective performance in open shelving?

## Methods

### Participants

Eleven older adults with MCI (mean age = 72.89 years, SD = 4.01; 6 males, 5 females; 8 White, 3 Black) participated in this study. Nine participants were recruited from a Cognitive Empowerment Program (Vickers et al., 2020), and two from an Alzheimer’s disease social program (Grill & Galvin, 2014). Inclusion criteria were an age of 65 years or older, proficiency in English, and either a clinical diagnosis of MCI or a Montreal Cognitive Assessment (MoCA) score ≤25 (mean = 21.63, SD = 3.06) (Nasreddine et al., 2005). All participants provided written informed consent prior to participation, and the study protocol received approval from the Institutional Review Board (IRB), with appropriate reliance agreements established with affiliated institutions.

### Research design and study setting

The study employed a within-subjects experimental design to compare meal preparation task performance under two conditions: closed cabinets (control) versus open shelving (experimental). To achieve open shelving, cabinet doors on one side of the symmetrical kitchen were detached. Block randomization assigned participants to condition order to minimize selection bias and learning effects (Suresh, 2011). The kitchen layout (Figure 1) and setup (Figure 2) were standardized across all participants. A 26-step recipe incorporating variant ingredients (detailed in Table 1) were used to reduce familiarity bias, and participants received a basic kitchen orientation before the experiment to further mitigate any influence from previous exposure or practice effects (Wang et al., 2022).

**Figure 1.**
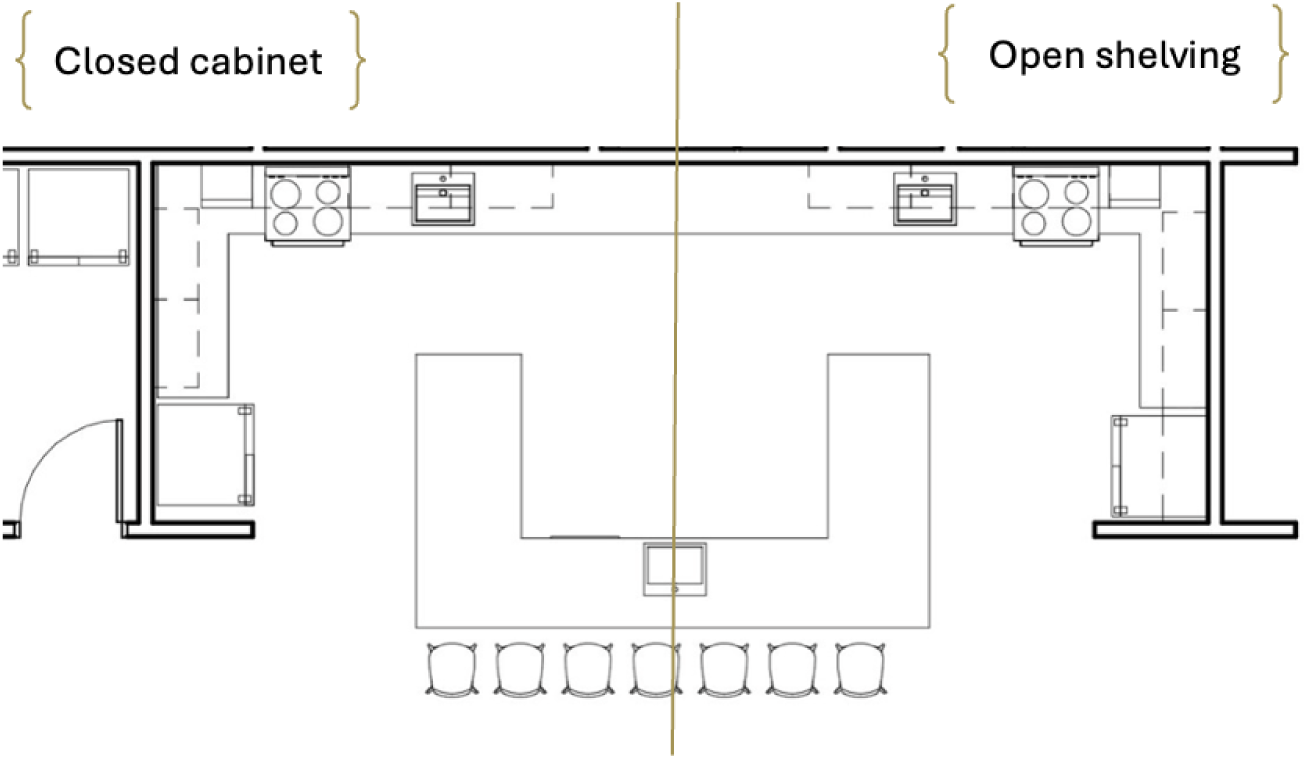
The kitchen layout of the study setting.

**Figure 2.**
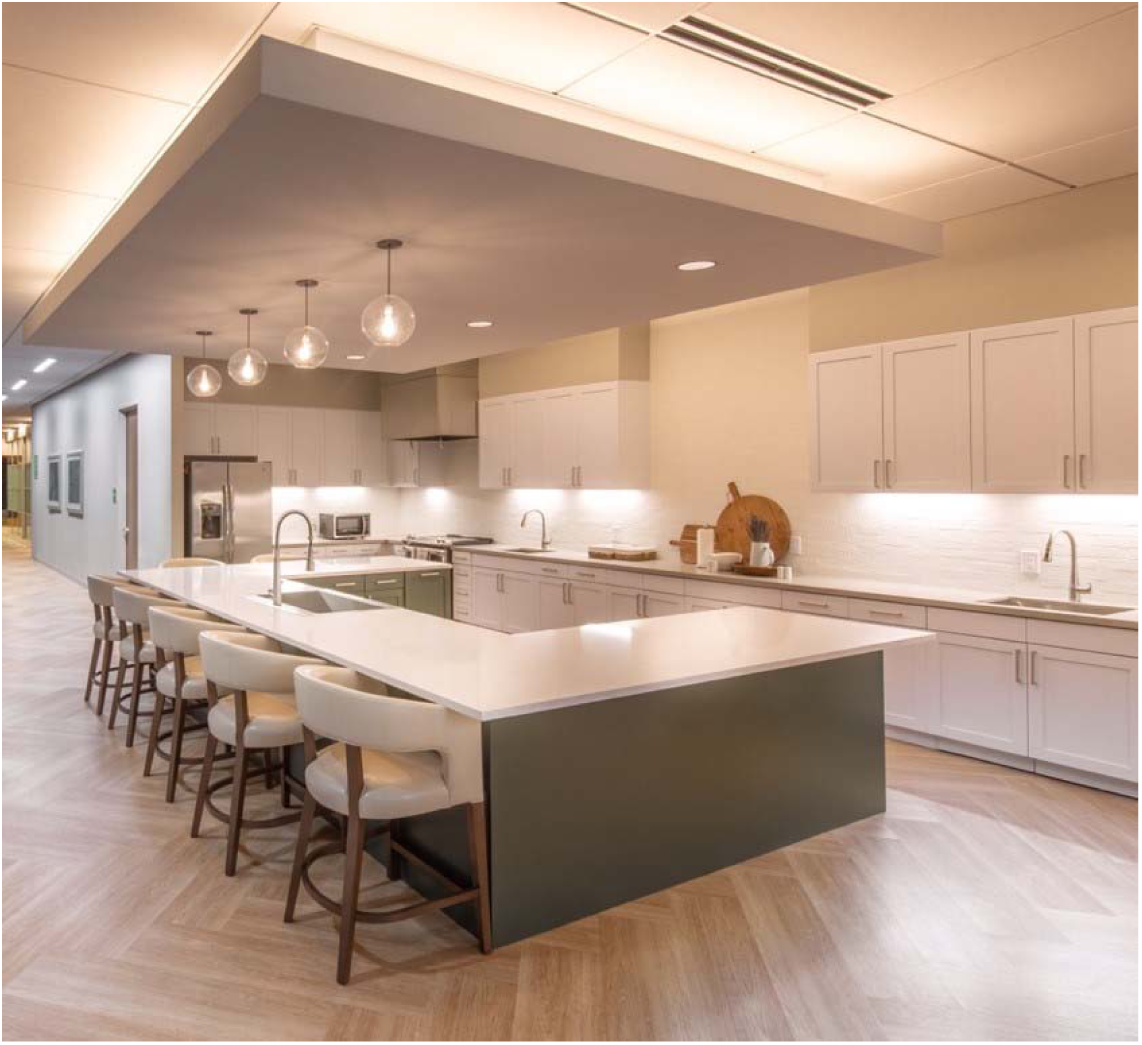
Image of the study setting.

**Table 1:**
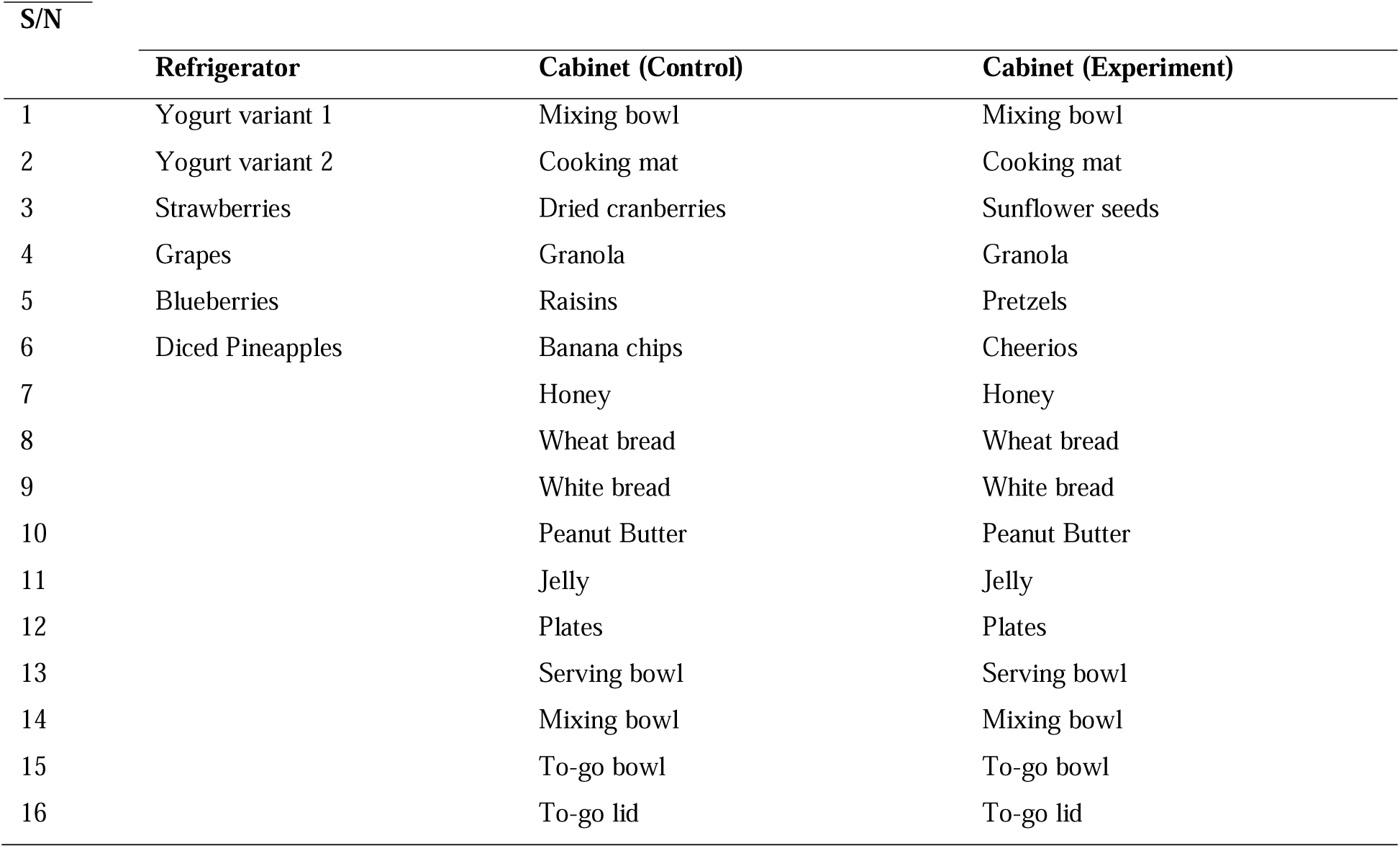
List and location of ingredients and utensils.

| S/N | Refrigerator | Cabinet (Control) | Cabinet (Experiment) |
| --- | --- | --- | --- |
| 1 | Yogurt variant 1 | Mixing bowl | Mixing bowl |
| 2 | Yogurt variant 2 | Cooking mat | Cooking mat |
| 3 | Strawberries | Dried cranberries | Sunflower seeds |
| 4 | Grapes | Granola | Granola |
| 5 | Blueberries | Raisins | Pretzels |
| 6 | Diced Pineapples | Banana chips | Cheerios |
| 7 |  | Honey | Honey |
| 8 |  | Wheat bread | Wheat bread |
| 9 |  | White bread | White bread |
| 10 |  | Peanut Butter | Peanut Butter |
| 11 |  | Jelly | Jelly |
| 12 |  | Plates | Plates |
| 13 |  | Serving bowl | Serving bowl |
| 14 |  | Mixing bowl | Mixing bowl |
| 15 |  | To-go bowl | To-go bowl |
| 16 |  | To-go lid | To-go lid |

### Measures

Cognitive load was assessed using the PupilLabs Core eye-tracking device (PupliLabs GmbH.), measuring fixation duration, fixation rate, saccade length, saccade velocity, pupil dilation, blink rate, and blink latency (Zagermann et al., 2016). Intrinsic motivation was measured using the Intrinsic Motivation Inventory (IMI; Interest/Enjoyment, Competence, and Pressure/Tension subscales) (Ryan, 1982). Physical activity intensity and step counts were recorded via GENEActiv wrist sensors (ActivInsights Ltd.), which measured acceleration magnitude (Freedson et al., 1998). Task duration was manually recorded using Excel or an online stopwatch (timeanddate.com). Additionally, two GoPro HERO9 cameras recorded participants’ performance throughout the experiment.

### Procedure

Participants randomly completed the assigned recipe under both conditions (experimental: open shelving, control: closed cabinets) while wearing eye trackers and wrist sensors. Data collection was conducted on weekdays from October 2023 to November 2024 between 2:00 and 5:00 p.m. EST. A detailed protocol ensured procedural consistency, including careful calibration of equipment and uniform placement of ingredients across conditions by trained researchers (Figures 3–5). A five-minute break and the completion of the IMI surveys followed each condition (detailed in Table 2).

**Figure 3.**
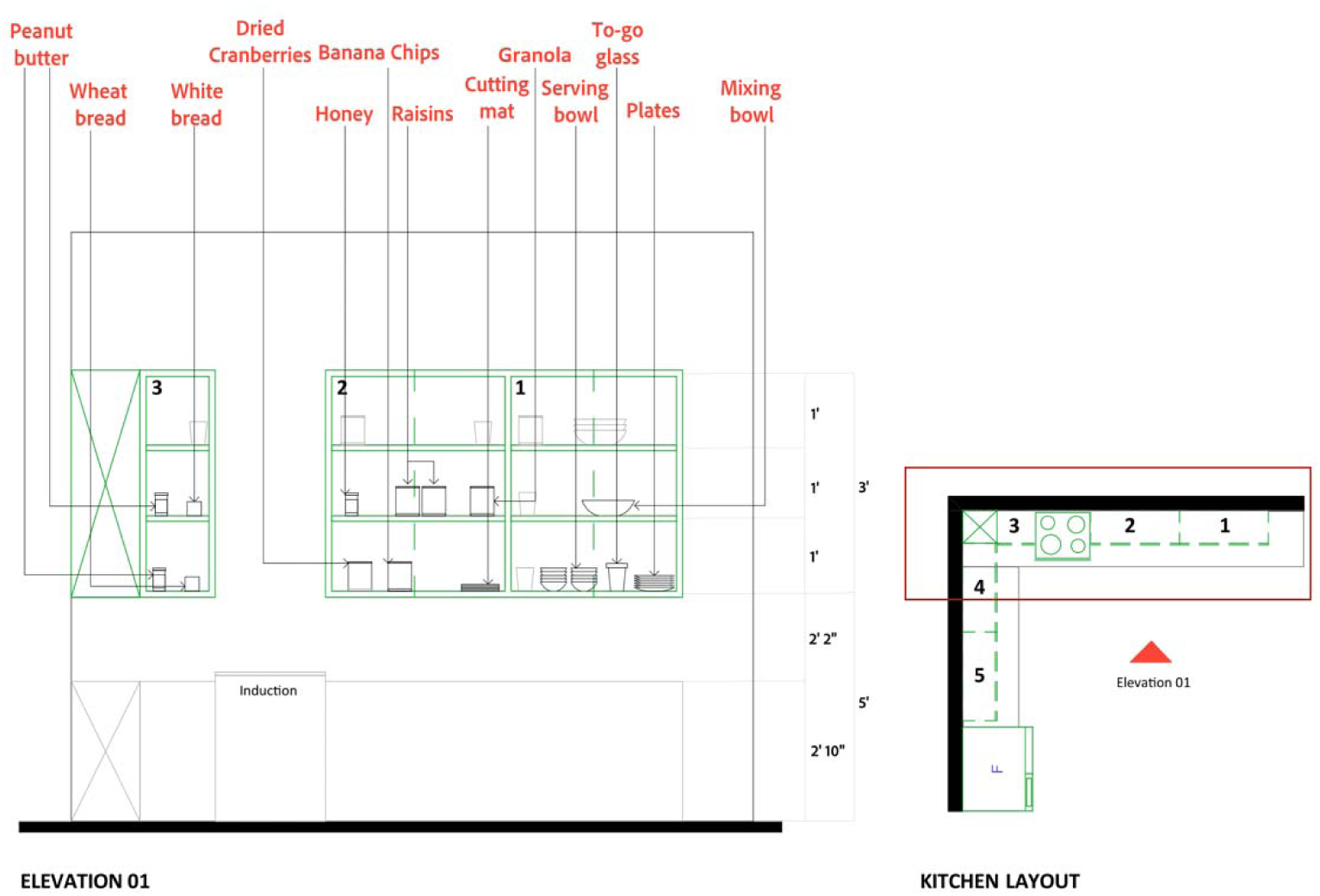
Placement of ingredients and utensils in the cabinet for control conditions.

**Figure 4.**
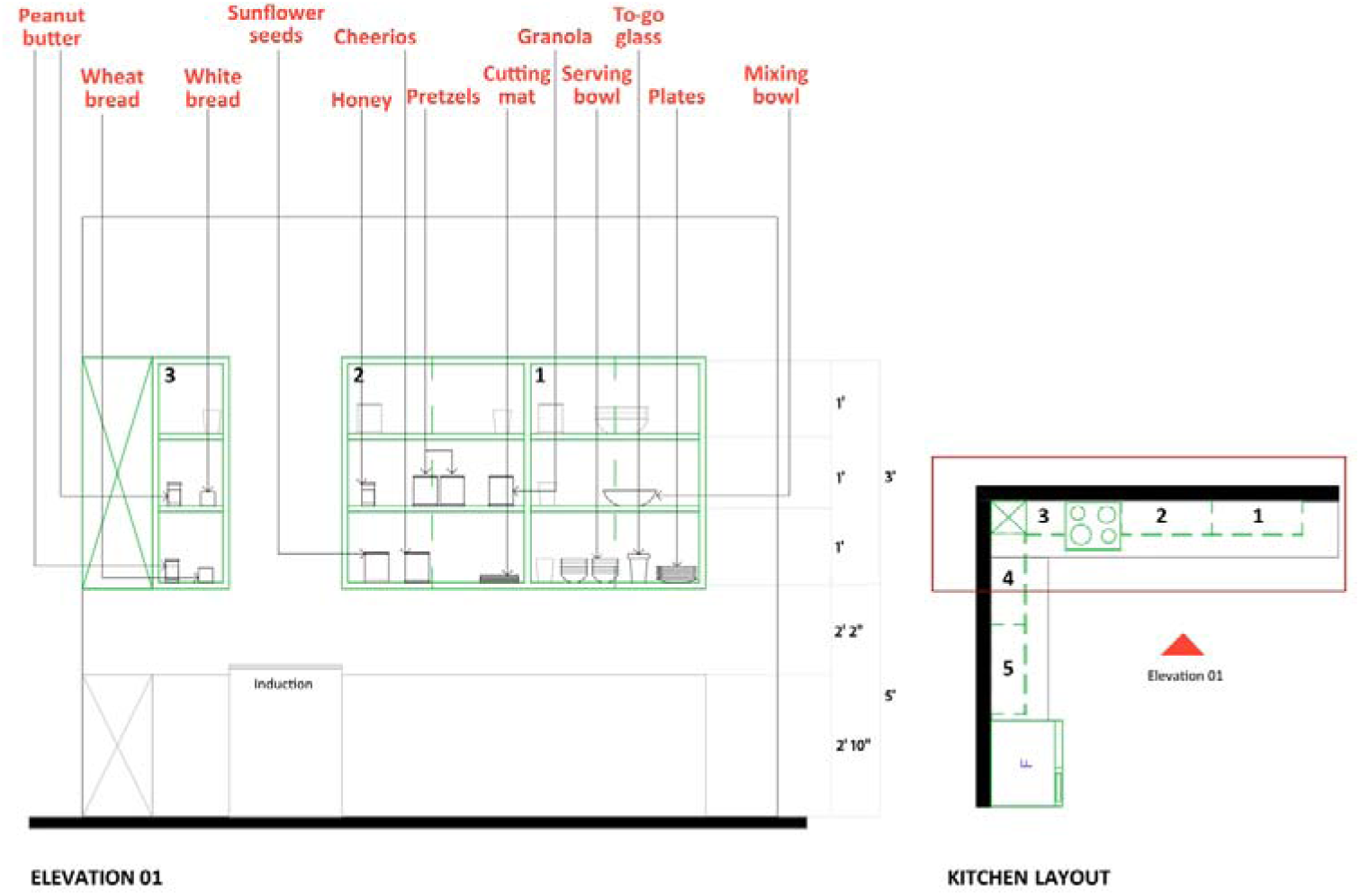
Placement of ingredients and utensils in the cabinet for experimental conditions.

**Figure 5.**
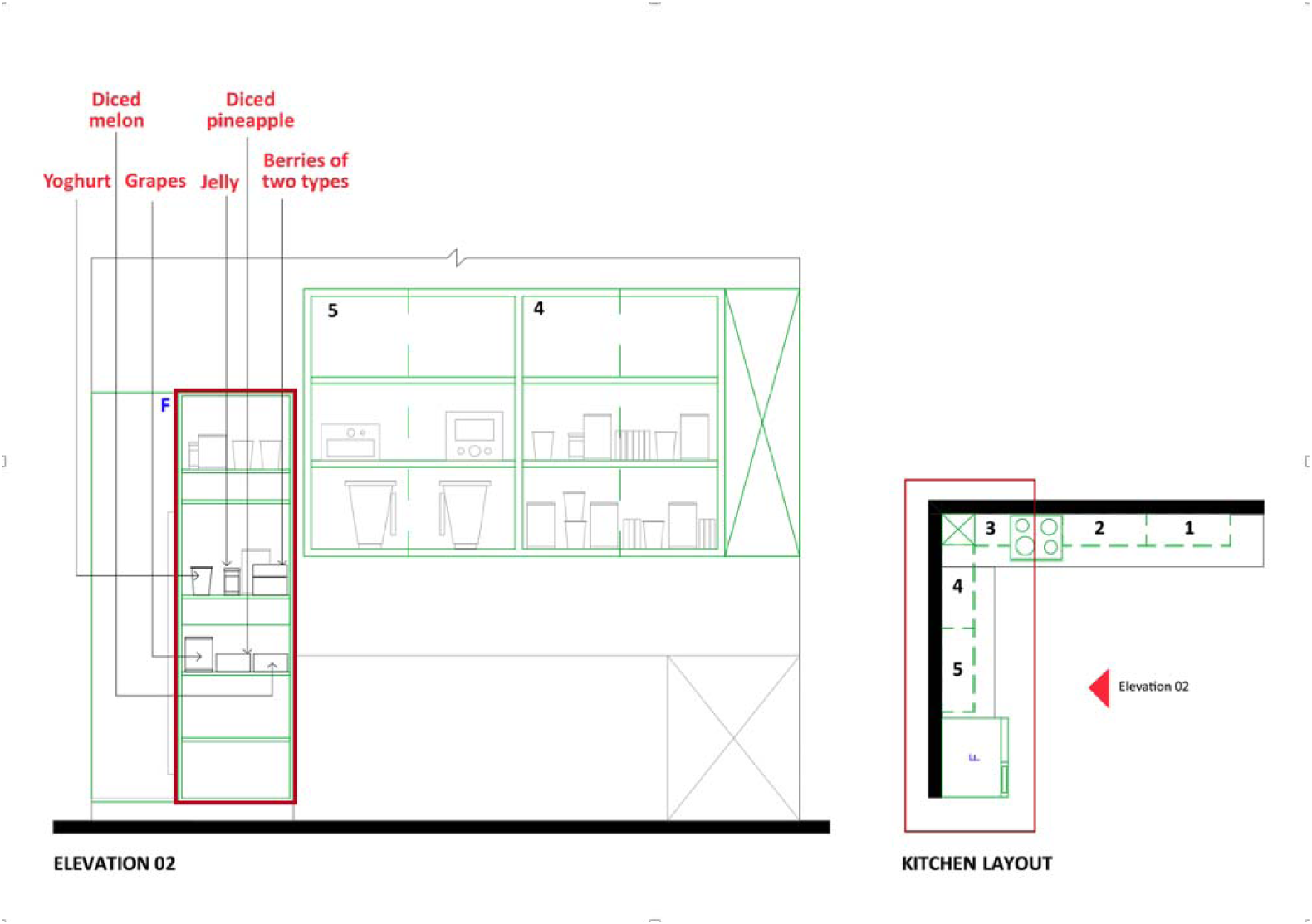
Placement of ingredients in the refrigerator for both control and experimental conditions.

**Table 2:** Eleven people within-subject design (the order of condition I and II exposure was randomized)

| Condition Order | Condition Type | Condition Details |
| --- | --- | --- |
| Condition 0 | Tutorial | Introducing the kitchen to the participants |
| *Preparation* | *Gadget setup* | *Participants put on the data collection devices (accelerometer and eye-tracker)* |
| Condition I | Control/Experiment | MCI participants are exposed to either the closed cabinets or open shelving based on randomization |
| *Break* | *For MCI participants* | *MCI participants take a minimum five-minute break, complete IMI survey and open needed interview on condition I* |
| Neutral Condition | Neutral | MCI participants are exposed to an unconnected but similar activity in the existing kitchen (closed cabinets) |
| *Break* | *For MCI participant* | *MCI participants take a break from the neutral condition* |
| Condition II | Control/Experiment | MCI participants are exposed to either the closed cabinet or open shelving based on randomization |
| *Break* | *For MCI participants* | *MCI participants complete IMI survey and open needed interview on condition II* |

### Data Analysis

Quantitative and qualitative data were collected from this study for data analysis, as shown in Table 3. Quantitative measures included eye-tracking metrics, wrist accelerometry, Intrinsic Motivation Inventory (IMI) subscales, and task duration. Data were cleaned and processed using Microsoft Excel, Python, and R software environments. IMI responses and task duration metrics were processed in Excel and then exported to R for statistical analysis and visualization. Eye-tracking and wrist sensor data were pre-processed in Python before final analyses were conducted in R.

**Table 3.** List of independent and dependent variables.

| Independent Variables (IVs) | Dependent Variables (DVs) |
| --- | --- |
| Visual accessibility (open shelving vs. closed cabinets)<br>(active IV) | Time duration (Time tracking) |
|  | Participants' motivation between both conditions (IMI scale) |
|  | Cognitive load (Eye-tracking-based measures) |
|  | Physical activity intensity and step count (Accelerometer-based measures) |

For eye-tracking data, metrics including fixation duration, fixation rate, saccade length, saccade velocity, pupil dilation, blink rate, and blink latency were extracted based on standard methods (Zagermann et al., 2016). Pupil dilation was calculated as the average of left and right pupil diameters, and saccades were identified using a velocity threshold method (>20°/s), following the procedures outlined by Salvucci and Goldberg (2000). It should be noted that due to equipment disconnections, complete eye-tracking data were unavailable for two participants (ID0004 and ID0007) in the control condition. This reflects known challenges associated with physiological monitoring technologies (Holmqvist et al., 2011).

For wrist accelerometry, gravitational acceleration was removed using a fourth-order Butterworth low-pass filter (cutoff: 0.3 Hz) (Anguita et al., 2013). Acceleration magnitude was then calculated as the root mean square of the three orthogonal axes following established conventions (Freedson et al., 1998).

IMI survey data were examined for missing values. In cases where limited missingness occurred (e.g., Pressure/Tension subscale for participant ID0006), mean imputation was employed, consistent with best practices for small-scale survey data (Ryan, 1982).

Descriptive statistics, including means, standard deviations, and 95% confidence intervals, were computed for each variable. Due to differing original scales across measures, z-score standardization was performed for participant profiling. Radar charts were constructed for each participant based on these z-scores to visually represent relative patterns across cognitive load, motivation, physical activity, and task duration.

Qualitative data were collected through post-task semi-structured interviews designed to explore participants’ perceptions of the open shelving condition. Interview transcripts were thematically analyzed using an inductive approach to identify recurrent patterns and themes, particularly relating to perceived benefits and barriers.

For exploratory participant profiling, z-scored variables were interpreted descriptively. Trends across cognitive load, motivation, physical activity, and task duration were used to classify participants into emerging typologies such as performers, strugglers, and motivated but physically strained, based on visual patterns observed in radar charts. Participants who consistently demonstrated improvements across all measures were termed “performers”. Participants who showed decline or minimal improvements across most measures were termed “strugglers”. Participants who showed increased motivation, but concurrently higher physical effort, were termed “motivated but physically strained”. Finally, participants who exhibited relatively stable z-scores near the sample mean were termed “balanced or average responders”.

### Statistical Analysis

All statistical analyses were performed using R (version 4.2.2) with base functions and packages, including stats, dplyr, and ggplot2. Data normality was assessed using the Shapiro-Wilk test for each variable to determine the appropriate inferential statistical tests (Royston, 1995). Variables that met normality assumptions, such as IMI subscales (Interest/Enjoyment, Perceived Competence, Pressure/Tension) and task duration, were analyzed using paired t-tests to compare performance between the closed cabinets (control) and open shelving (experimental) conditions.

Conversely, cognitive load measures, such as fixation rate, fixation duration, saccade length, blink rate, and blink latency, were found to violate normality assumptions (p < 0.05) according to Shapiro-Wilk tests. Consequently, the non-parametric Wilcoxon signed-rank test was employed for these variables (Hollander et al., 2013). Similarly, physical activity measures derived from wrist accelerometry (acceleration magnitude and step count) exhibited non-normal distributions, as confirmed by the Anderson-Darling and Lilliefors tests (Stephens, 1974), and were analyzed using Wilcoxon signed-rank tests.

The small sample size limited statistical power, necessitating a focus on effect sizes and confidence intervals rather than strict reliance on significance testing (Lakens, 2013). Effect sizes were calculated using Cohen’s dz for paired t-tests and matched rank-biserial correlations for Wilcoxon tests, following guidelines for small-sample designs (Lakens, 2013). Ninety-five percent confidence intervals were reported for both effect sizes and mean differences to supplement p-values, following recommendations to prioritize estimation over significance testing in the context of pilot studies (Sullivan & Feinn, 2012).

## Results

### Group-Level Outcomes

#### Cognitive Load

Analysis of eye-tracking metrics showed mixed effects regarding the impact of shelving type. Mean fixation rate increased slightly in the open shelving compared to closed cabinets condition (dz = -0.04), while blink latency decreased moderately (dz = 0.32), suggesting possible improvements in visual processing speed with open shelving. Fixation rate and blink rate demonstrated minimal differences between the two conditions.

**Figure 6.**
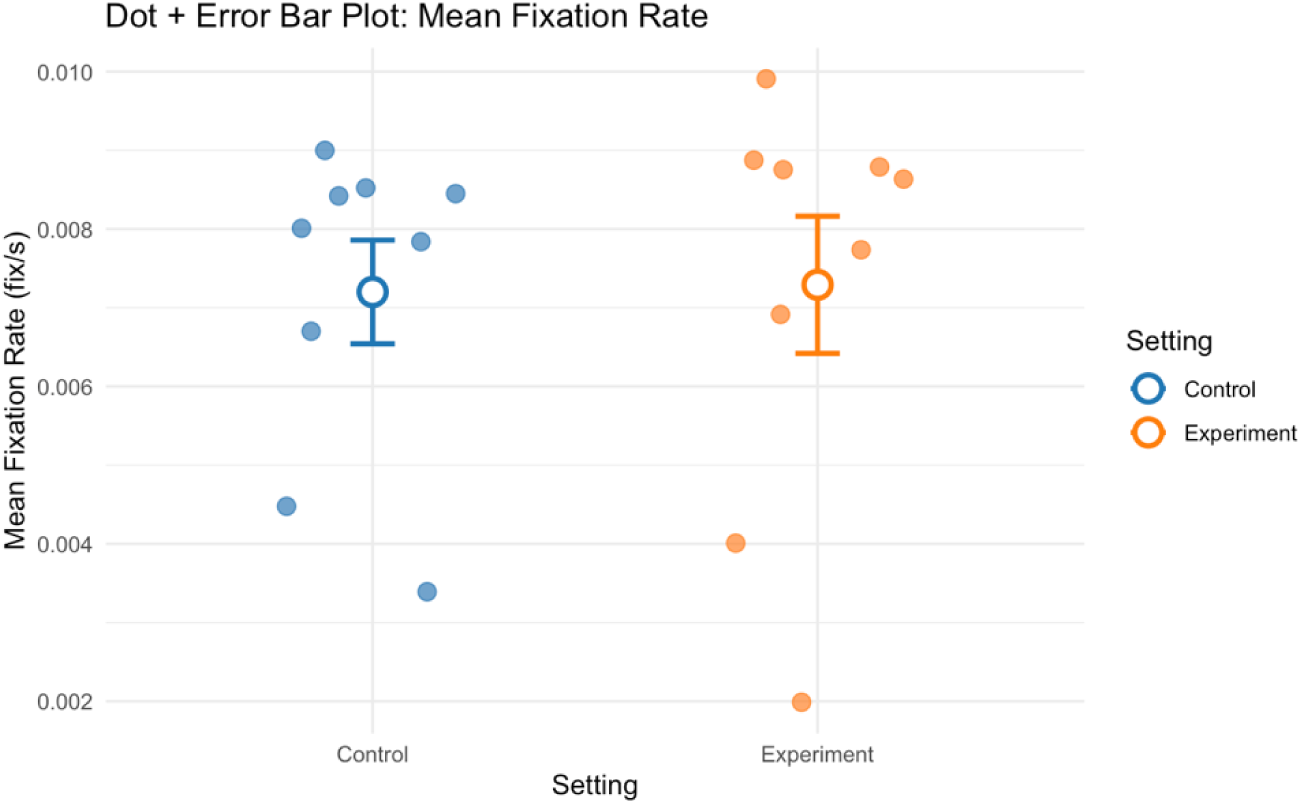
Group mean fixation rate for all participants in control and experiment conditions.

**Figure 7.**
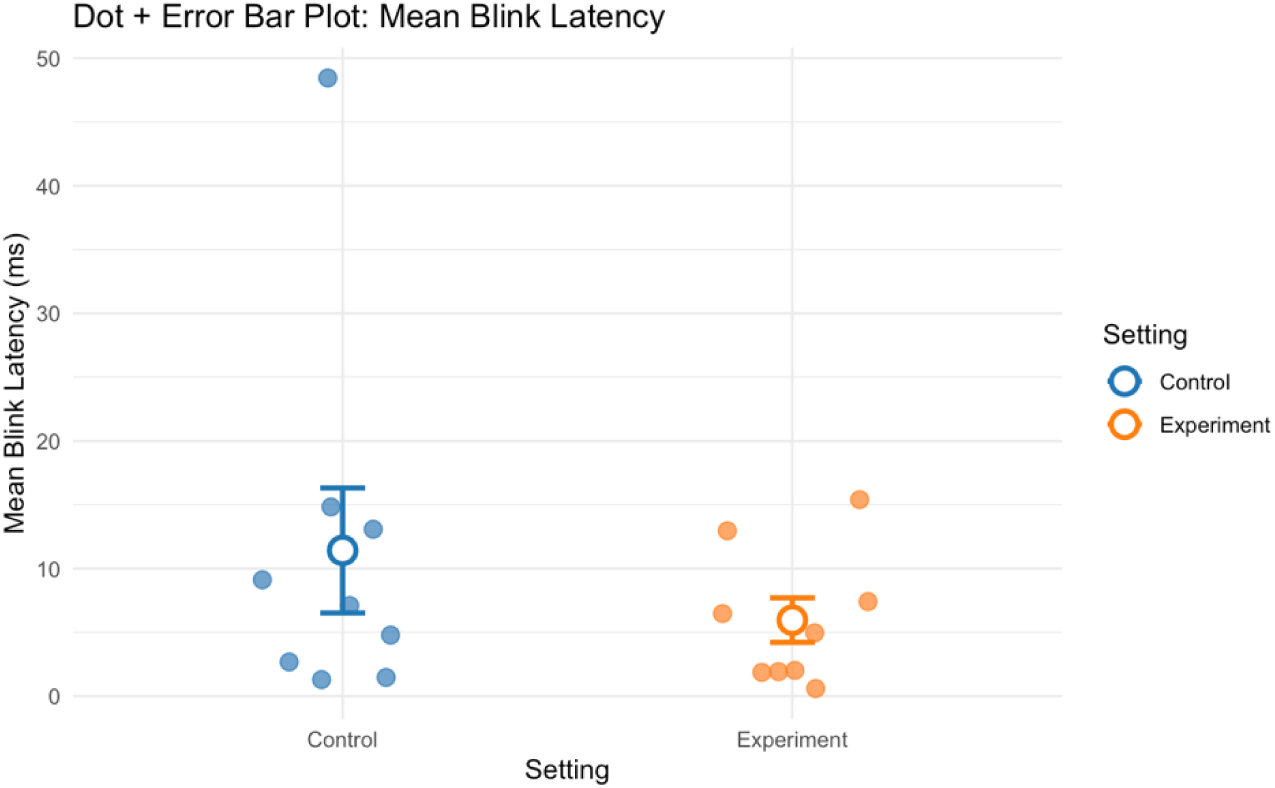
Group mean blink latency for all participants in control and experiment conditions.

#### Intrinsic Motivation

IMI survey results indicated that participants reported higher perceived competence (dz = 0.56) and lower pressure/tension (dz = -0.49) under the open shelving condition compared to the closed cabinets condition, suggesting enhanced intrinsic motivation associated with open shelving. Interest/enjoyment also trended upward (dz = 0.32).

**Figure 8.**
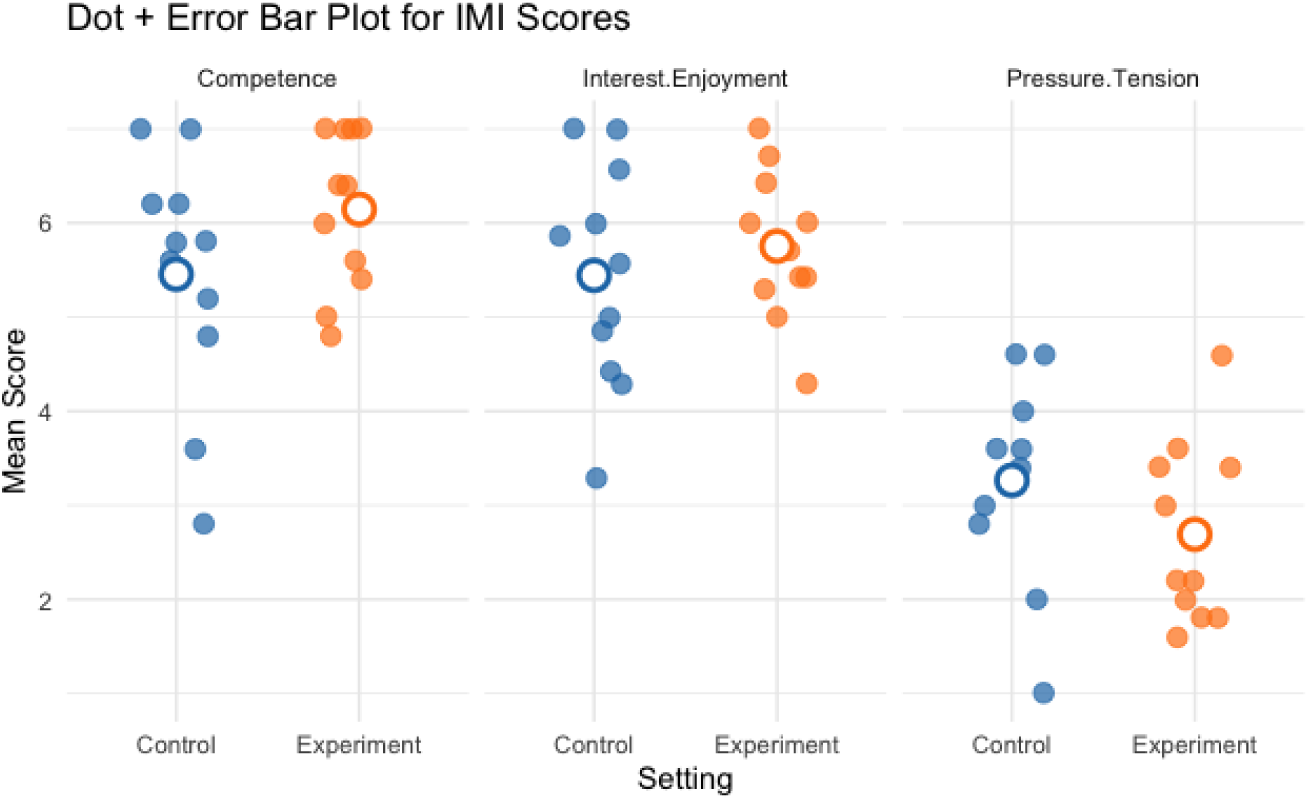
Group mean IMI scores (perceived competence, interest/enjoyment & pressure/tension) for all participants in control and experiment conditions.

#### Physical Activity Intensity

Regarding physical exertion, mean acceleration magnitude exhibited a slight yet statistically significant decrease in the open shelving condition compared to the closed cabinets condition (dz = 0.06; p < 0.001). This finding indicates a minor reduction in overall physical effort when participants interacted with the open shelving configuration.

**Figure 9.**
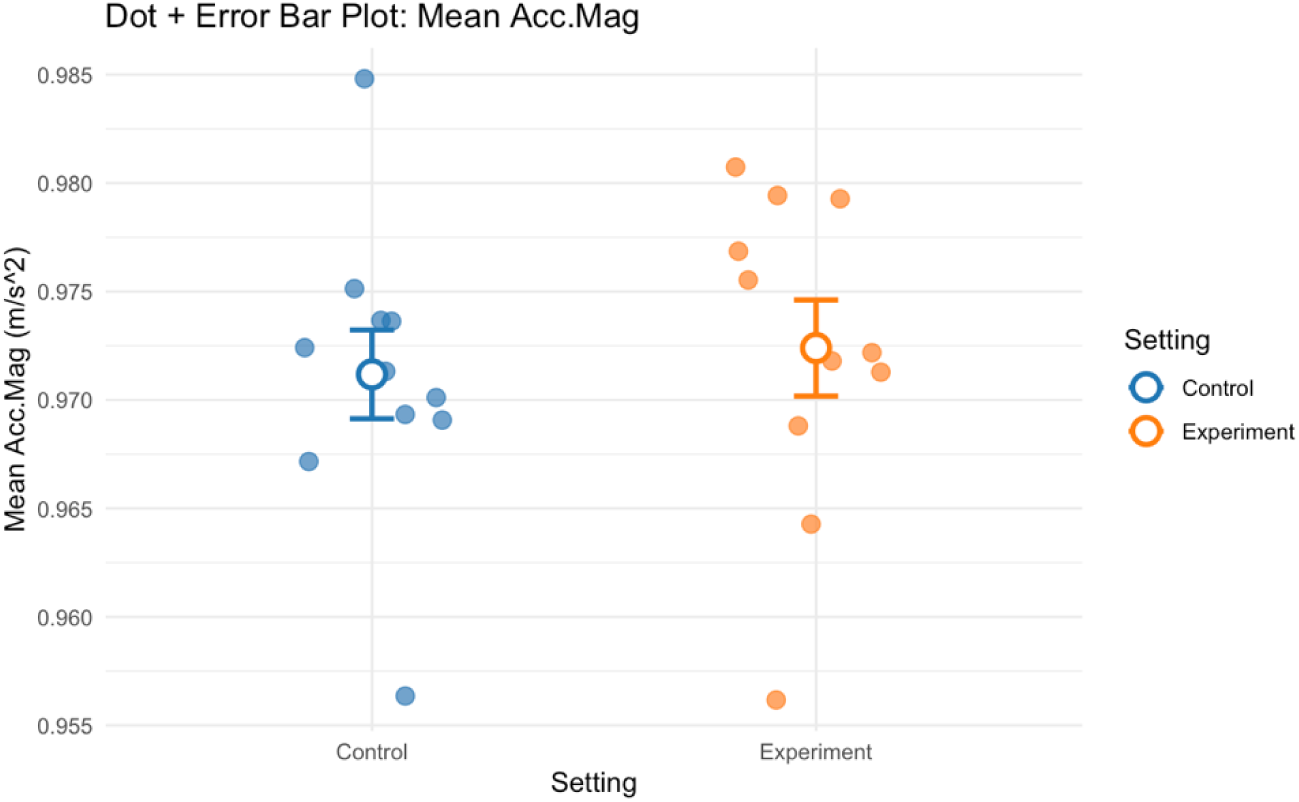
Group mean acceleration magnitude for all participants in control and experiment conditions.

#### Step Count

Step count showed a substantial decrease from the closed cabinet to open shelving settings. The Wilcoxon signed-rank test indicated this reduction was statistically significant with a large effect size (dz = 0.66; p = 0.002). This suggests that open shelving notably reduces unnecessary movement during kitchen tasks.

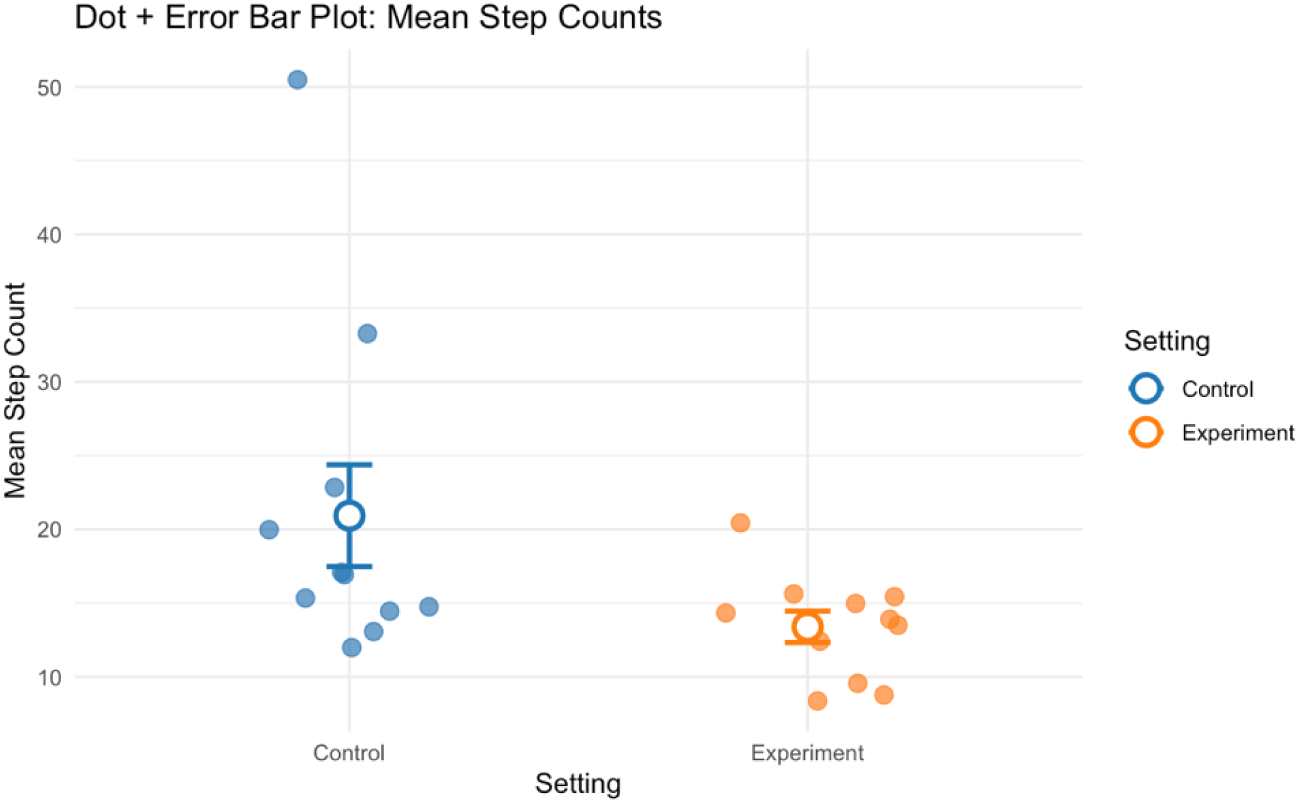

#### Task Duration

Mean task duration was slightly higher in closed cabinets than in open shelving conditions (dz = -0.56), suggesting increased efficiency with open shelving.

### Individual-Level Participant Profiles

#### Quantitative Profiles and Radar Charts

Distinct participant typologies emerged from these profiles based on the z-score performance across all measures. For instance, some participants (i.e., ID0005 and ID0011) fell under the “performers” group. Other participants (i.e., ID0003 and ID0006) fell under “strugglers.” A subset of participants, such as ID0002, fell under the “motivated but physically strained” group. Finally, participants such as ID0008 and ID0009 fell under the “balanced or average responders” group.

Table 5 presents a summary of each participant’s demographic profile, cognitive status, trends across measured variables, and overall classification. Selected radar charts (Figures 11–17) illustrate representative examples of the different participant response types. Full radar charts for all eleven participants are available in the supplementary materials.

**Figure 10.**
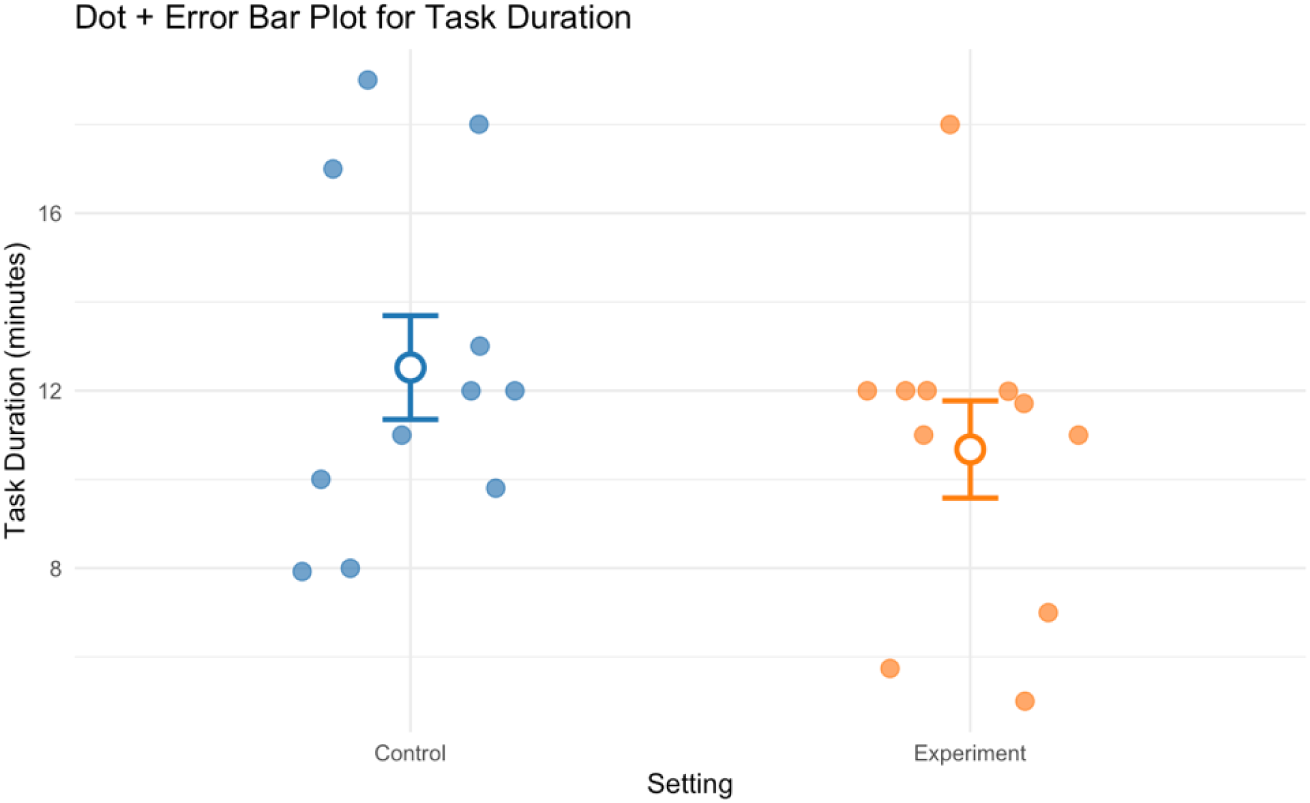
Group mean task duration for all participants in control and experiment conditions.

**Figure 11.**
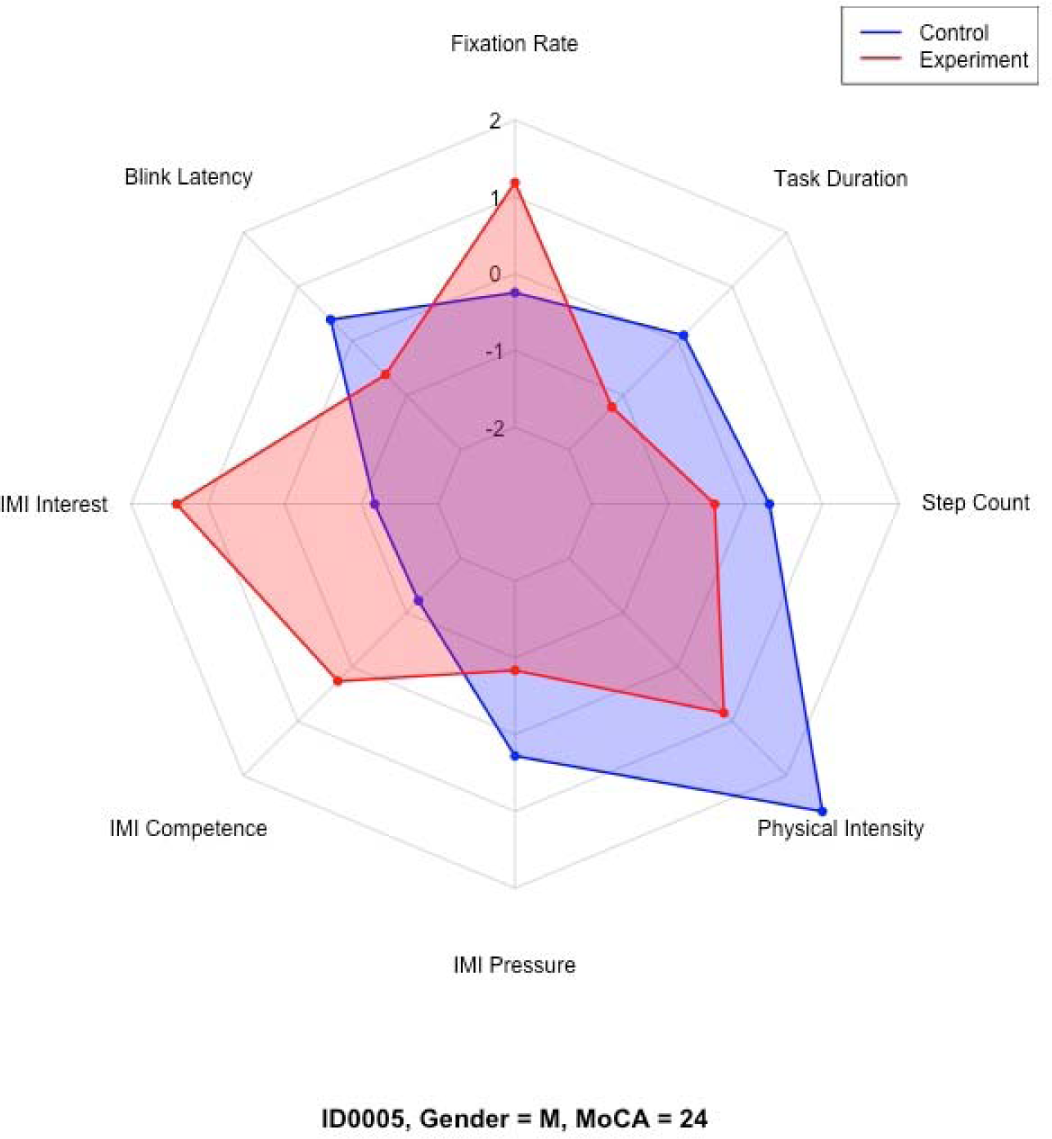
Representative radar charts of the “performers” participant group.

**Figure 12.**
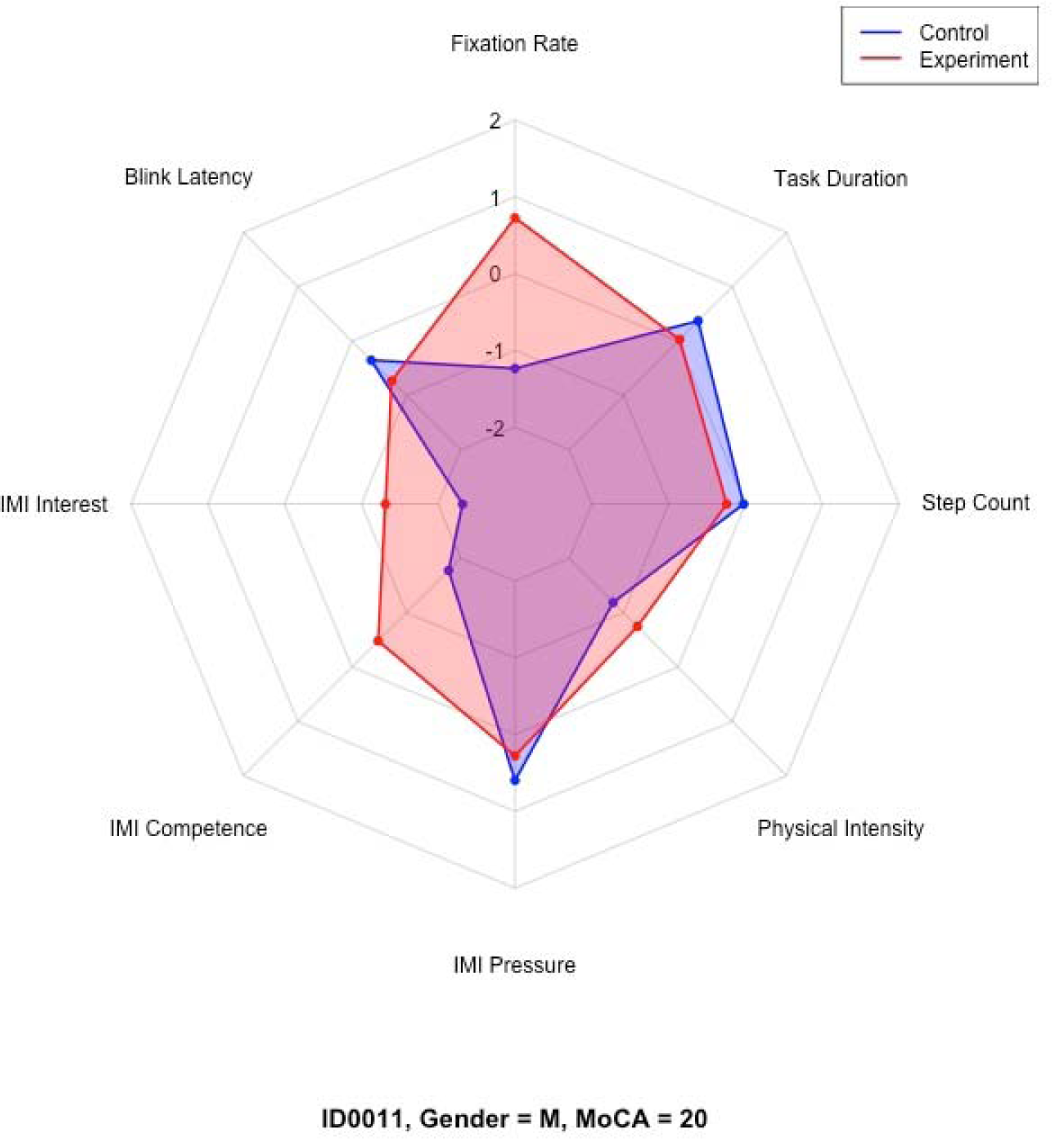
Representative radar charts of the “performers” participant group.

**Figure 13.**
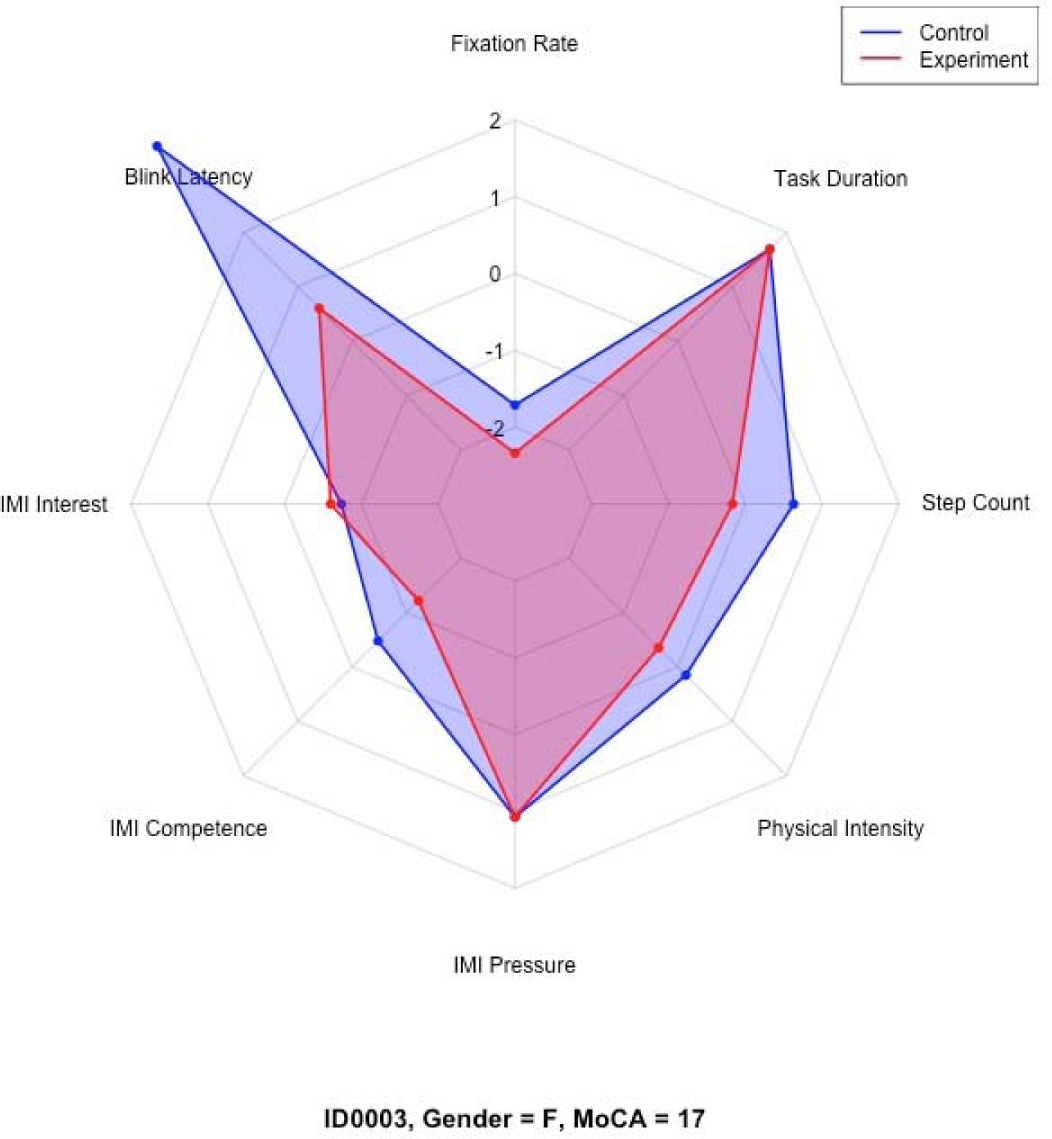
Representative radar charts of the “strugglers” participant group.

**Figure 14.**
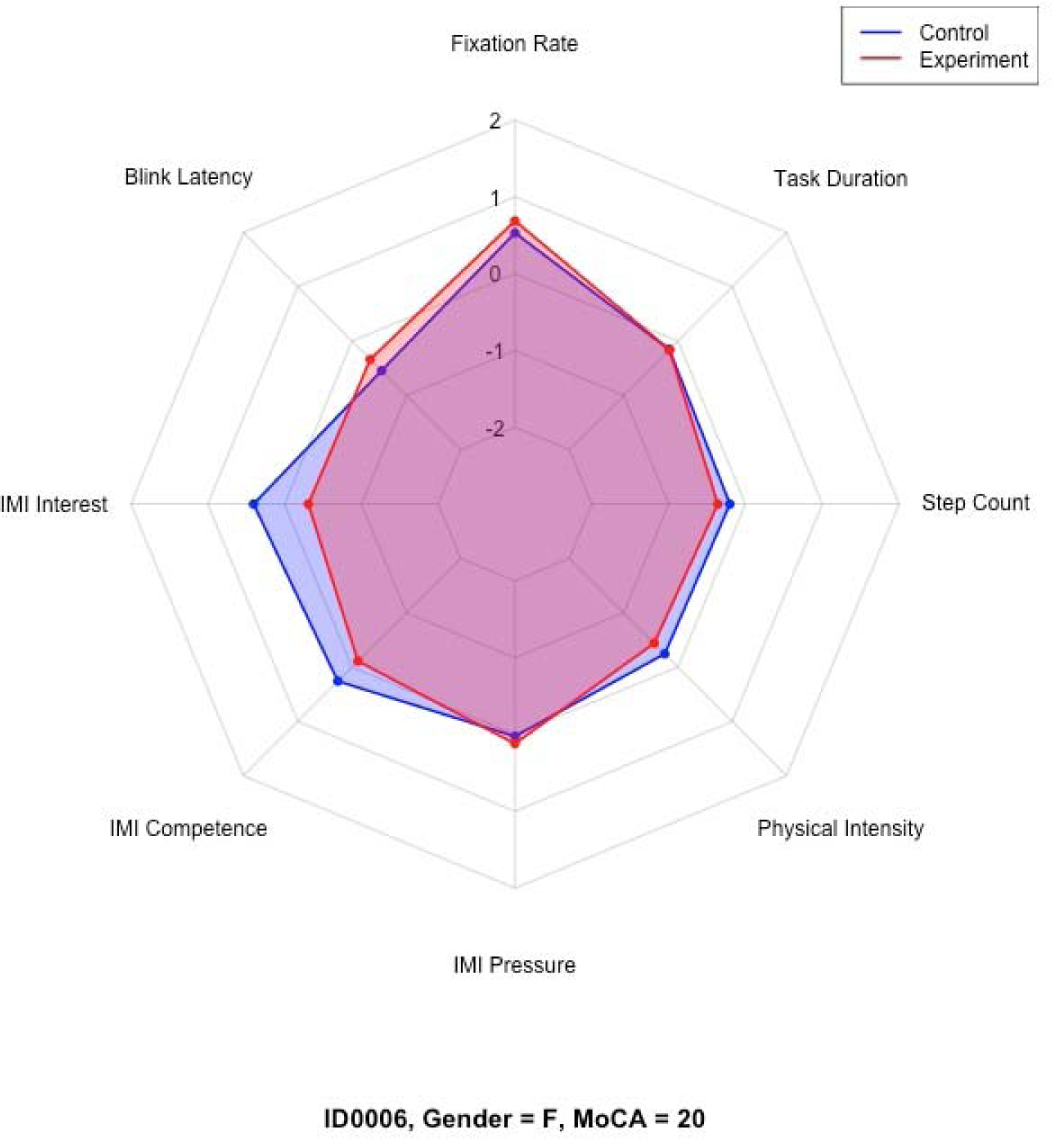
Representative radar charts of the “strugglers” participant group.

**Figure 15.**
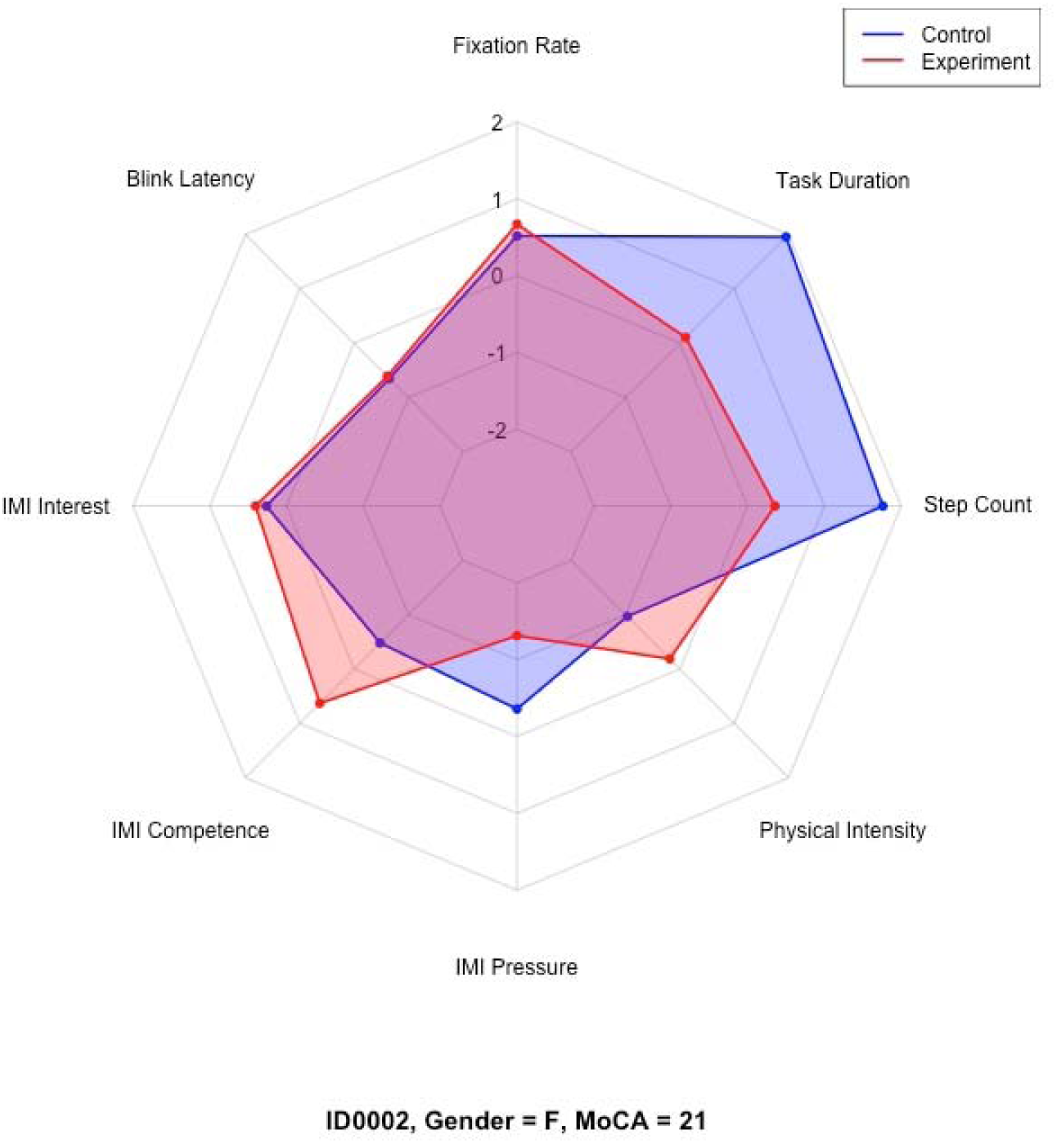
Representative radar charts of the “motivated but physically strained” participant group.

**Figure 16.**
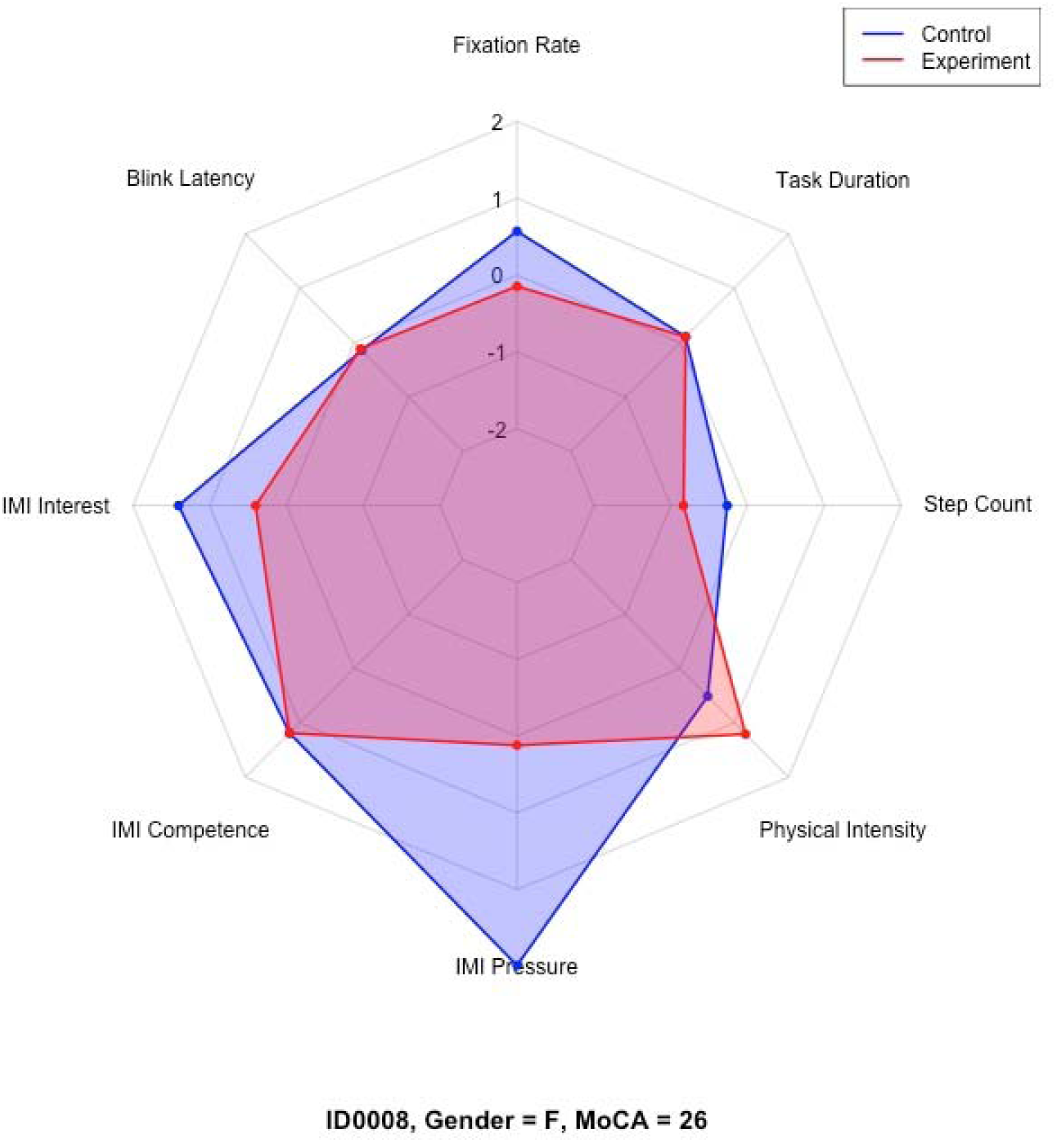
Representative radar charts of the “balanced or average responders” participant group.

**Figure 17.**
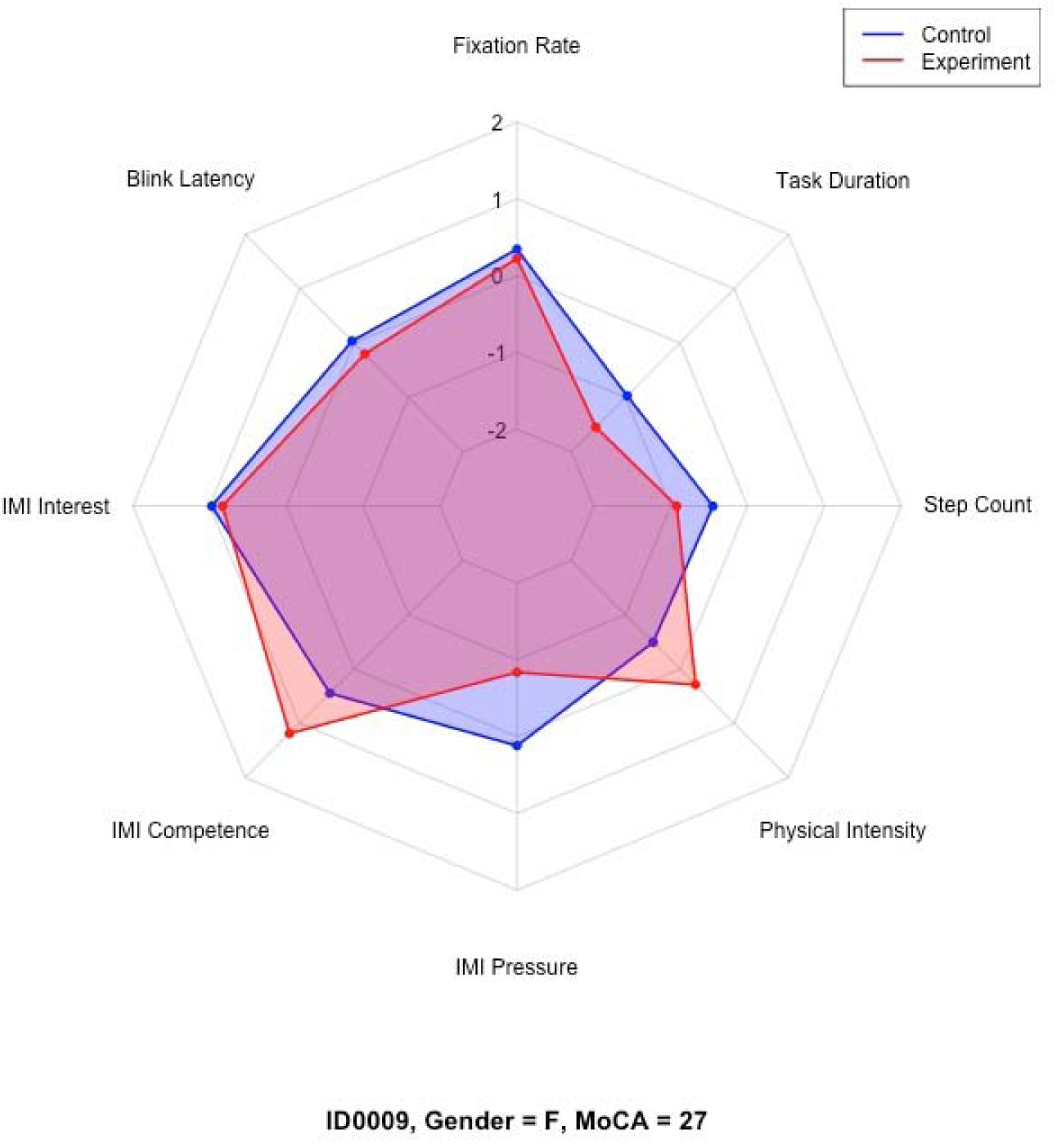
Representative radar charts of the “balanced or average responders” participant group.

**Table 4.**
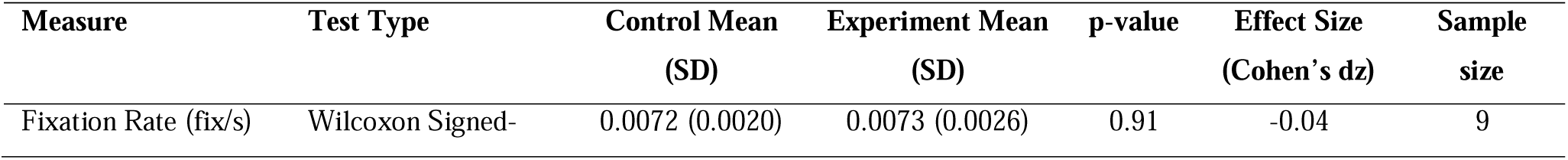
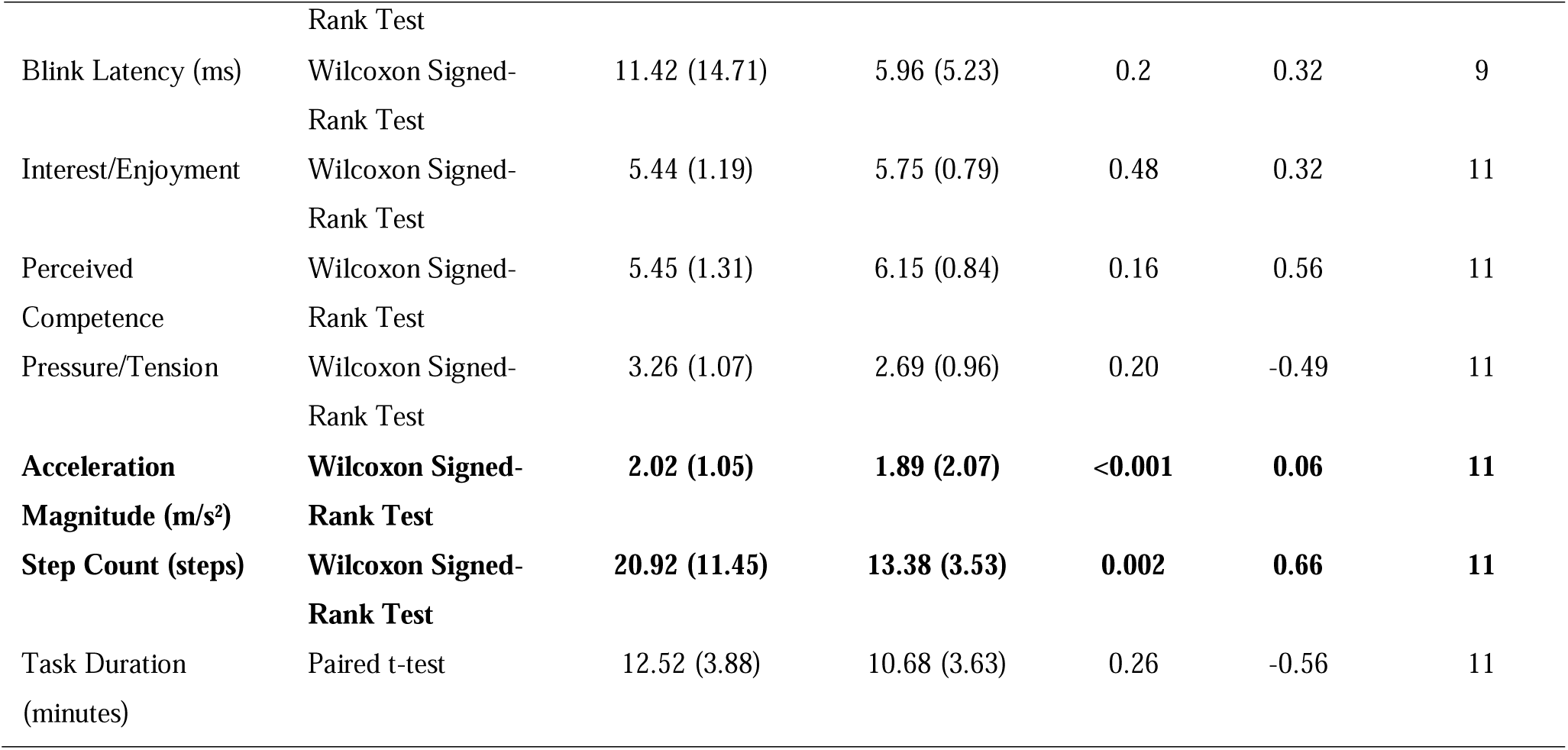
Summary of Group Comparisons across Key Measures.

**Table 5.** Summary of each participant’s demographic profile, cognitive status, trends across measured variables, and quantitative profile.

| ID | Gender | MoCA | Quantitative Profile | Fixation Rate (fix/s) (Mean) | Blink Latency (ms) (Mean, CI) | IMI Interest/Enjoyment (Mean, CI) | IMI Perceived Competence (Mean, CI) | IMI Pressure/Tension (Mean, CI) | Physical Activity Intensity - Acc.Mag (m/s ^2) (Mean, CI) | Step Count (Mean, CI) | Task Duration (min) (Mean) | Trends (C→E) |
| --- | --- | --- | --- | --- | --- | --- | --- | --- | --- | --- | --- | --- |
| 0001 | Female | 20 | Balanced or average responder | C: 0.00899;<br>E: 0.00863 | C: 1.277<br>[0.906, 1.648];<br>E: 2.025<br>[1.249, 2.801] | C: 5.00 [4.96,<br>5.04]; E: 5.43<br>[5.38, 5.47] | C: 6.60 [6.58, 6.63];<br>E: 6.80 [6.79, 6.81] | C: 2.00 [1.95, 2.05];<br>E: 1.80 [1.77, 1.83] | C: 2.01<br>[2.00, 2.02]; E:<br>1.64 [1.63, 1.64] | C: 17.06<br>[9.34, 24.78]; E:<br>15.44<br>[10.72, 20.16] | C: 8; E:<br>11 | ↓ FixRate, ↑ BliRate, ↑ IMI-Int, ↑ IMI-Comp, ↓ IMI-Press, ↓ Acc, ↓ Step, ↑ Time |
| 0002 | Female | 21 | Motivated but physically strained | C: 0.00841;<br>E: 0.00875 | C: 1.482<br>[1.097, 1.866];<br>E: 1.916<br>[1.308, 2.525] | C: 5.86 [5.81,<br>5.91]; E: 6.00<br>[5.98, 6.02] | C: 5.20 [5.13, 5.27];<br>E: 6.40 [6.39, 6.42] | C: 2.80 [2.75, 2.85];<br>E: 1.60 [1.58, 1.63] | C: 1.69<br>[1.68, 1.70]; E:<br>1.97 [1.96, 1.97] | C: 33.25<br>[22.57, 43.93]; E:<br>20.44<br>[13.80, 27.08] | C: 19; E:<br>12 | ↑ FixRate, ↑ BliRate, ↑ IMI-Int, ↑ IMI-Comp, ↓ IMI-Press, ↑ Acc, ↓ Step, ↓ Time |
| 0003 | Female | 17 | Struggler | C: 0.00340;<br>E: 0.00199 | C: 48.440 [-<br>20.517, 117.396]; E:<br>15.398 [6.375, 24.421] | C: 4.86 [4.83,<br>4.89]; E: 5.00<br>[4.95, 5.05] | C: 5.20 [5.17, 5.23];<br>E: 4.40 [4.35, 4.45] | C: 4.60 [4.55, 4.65];<br>E: 4.60 [4.54, 4.66] | C: 2.09<br>[2.09, 2.10]; E:<br>1.91 [1.91, 1.92] | C: 22.88<br>[16.31, 29.44]; E:<br>15.62<br>[11.98, 19.27] | C: 18; E:<br>18 | ↓ FixRate, ↑ BliRate, ↓ IMI-Int, ↓ IMI-Comp, ≈ IMI-Press, ↓ Acc, ↓ Step, ≈ Time |
| 0004 | Female | 21 | Balanced or average responder | C: NA; E:<br>NA | C: NA; E: NA | C: 7.00 [NA]; E:<br>6.71 [6.70, 6.73] | C: 6.40 [6.39, 6.42];<br>E: 6.60 [6.59, 6.62] | C: 4.60 [4.56, 4.64];<br>E: 3.00 [2.95, 3.05] | C: 2.15<br>[2.14, 2.16]; E:<br>2.27 [2.26, 2.28] | C: 50.44<br>[40.78, 60.09]; E:<br>14.31<br>[9.29, 19.34] | C: 17; E:<br>12 | ↓ IMI-Int, ↑ IMI-Comp, ↓ IMI-Press, ↑ Acc, ↓ Step, ↓ Time |
| 0005 | Male | 24 | Performer | C: 0.00669;<br>E: 0.00991 | C: 13.064<br>[5.977, 20.151]; E:<br>1.864 [0.951, 2.778] | C: 4.43 [4.37,<br>4.49]; E: 7.00 [NA] | C: 4.40 [4.33, 4.47];<br>E: 6.00 [NA] | C: 3.60 [3.53, 3.67];<br>E: 2.20 [2.13, 2.28] | C: 2.99<br>[2.98, 3.01]; E:<br>2.34 [2.32, 2.35] | C: 20.00<br>[13.11, 26.89]; E:<br>13.50<br>[6.51, 20.49] | C: 12; E:<br>7 | ↑ FixRate, ↑ BliRate, ↓ IMI-Int, ↑ IMI-Comp, ↓ IMI-Press, ↓ Acc, ↓ Step, ↓ Time |
| 0006 | Female | 20 | Struggler | C: 0.00844;<br>E: 0.00879 | C: 2.679<br>[1.884, 3.474];<br>E: 4.973<br>[3.395, 6.551] | C: 6.00 [5.98,<br>6.02]; E: 5.29<br>[5.25, 5.32] | C: 6.00 [NA]; E:<br>5.60 [5.59, 5.62] | C: 3.28 [3.25, 3.31];<br>E: 3.40 [3.36, 3.44] | C: 1.95<br>[1.95, 1.96]; E:<br>1.88 [1.87, 1.89] | C: 15.31<br>[12.79, 17.83]; E:<br>13.88<br>[10.29, 17.46] | C: 11; E:<br>11 | ↑ FixRate, ↓ BliRate, ↑ IMI-Int, ↓ IMI-Comp, ↑ IMI-Press, ↓ Acc, ↓ Step, ≈ Time |
| 0007 | Male | 25 | Performer | C: NA; E:<br>NA | C: NA; E: NA | C: 4.29 [4.26,<br>4.31]; E: 5.43<br>[5.40, 5.46] | C: 3.00 [NA]; E:<br>5.80 [5.78, 5.82] | C: 3.60 [3.56, 3.64];<br>E: 1.80 [1.78, 1.82] | C: 1.78<br>[1.77, 1.79]; E:<br>1.46 [1.45, 1.47] | C: 14.44<br>[11.59, 17.28]; E:<br>8.38<br>[6.07, 10.68] | C: 10; E:<br>5 | ↑ IMI-Int, ↑ IMI-Comp, ↓ IMI-Press, ↓ Acc, ↓ Step, ↓ Time |
| 0008 | Female | 26 | Balanced or average responder | C: 0.00853;<br>E: 0.00692 | C: 7.116<br>[4.710, 9.522];<br>E: 7.410<br>[5.155, 9.664] | C: 7.00 [NA]; E:<br>6.00 [5.96, 6.05] | C: 7.00 [NA]; E:<br>7.00 [NA] | C: 7.00 [NA]; E:<br>3.40 [3.32, 3.48] | C: 2.22<br>[2.21, 2.23]; E:<br>2.47 [2.45, 2.48] | C: 14.75<br>[12.31, 17.19]; E:<br>9.56<br>[6.17, 12.96] | C: 12; E:<br>12 | ↓ FixRate, ↓ BliRate, ↑ IMI-Int, ≈ IMI-Comp, ↓ IMI-Press, ↑ Acc, ↓ Step, ≈ Time |
| 0009 | Female | 27 | Balanced or average responder | C: 0.00802;<br>E: 0.00775 | C: 9.104<br>[2.857,<br>15.351]; E:<br>6.492 [2.308,<br>10.677] | C: 6.57 [6.55,<br>6.59]; E: 6.43<br>[6.42, 6.44] | C: 6.20 [6.18, 6.22];<br>E: 7.00 [NA] | C: 3.40 [3.35, 3.45];<br>E: 2.20 [2.13, 2.28] | C: 1.86<br>[1.85,<br>1.87]; E:<br>2.14 [2.12,<br>2.15] | C: 13.06<br>[9.75,<br>16.38]; E:<br>8.75<br>[6.73,<br>10.77] | C: 7.92;<br>E: 5.74 | ↓ FixRate, ↓<br>BliRate, ↓ IMI-Int, ↑ IMI-Comp, ↓ IMI-Press, ↑ Acc, ↓<br>Step, ↓ Time |
| 0010 | Male | 23 | Balanced or average responder | C: 0.00785;<br>E: 0.00401 | C: 14.841<br>[4.569,<br>25.112]; E:<br>12.958 [5.599,<br>20.318] | C: 5.57 [5.54,<br>5.60]; E: 5.71<br>[5.67, 5.76] | C: 5.80 [5.78, 5.82];<br>E: 5.40 [5.36, 5.44] | C: 3.00 [2.92, 3.08];<br>E: 2.00 [1.96, 2.04] | C: 1.96<br>[1.95,<br>1.97]; E:<br>2.65 [2.63,<br>2.67] | C: 12.00<br>[7.99,<br>16.01]; E:<br>12.40<br>[8.61,<br>16.19] | C: 9.8; E:<br>11.99 | ↓ FixRate, ↓<br>BliRate, ↓ IMI-Int, ↓ IMI-Comp, ↓ IMI-Press, ↑ Acc, ↑<br>Step, ↑ Time |
| 0011 | Male | 20 | Performer | C: 0.00447;<br>E: 0.00888 | C: 4.788<br>[3.504, 6.073];<br>E: 0.603<br>[0.367, 0.840] | C: 3.29 [3.25,<br>3.32]; E: 4.29<br>[4.26, 4.31] | C: 3.80 [3.78, 3.82];<br>E: 5.20 [5.15, 5.25] | C: 4.00 [3.93, 4.07];<br>E: 3.60 [3.55, 3.65] | C: 1.61<br>[1.60,<br>1.61]; E:<br>1.77 [1.76,<br>1.78] | C: 16.94<br>[9.33,<br>24.55]; E:<br>14.93<br>[8.11,<br>21.75] | C: 13; E:<br>11.71 | ↑ FixRate, ↑<br>BliRate, ↓ IMI-Int, ↑ IMI-Comp, ↓ IMI-Press, ↑ Acc, ↓<br>Step, ↓ Time |

*Note: C = Control, E = Experiment. Trends indicate direction of change from Control to Experiment (↑ = increase, ↓ = decrease, ≈ = no clear change). All measures include 95% confidence intervals where available. NA indicates missing data.*

#### Qualitative Perceptions of Open Shelving

Post-task semi-structured interviews revealed a range of participant perceptions regarding open shelving intervention. Participants who expressed positive sentiments, such as ID0005, ID0007, ID0009, and ID0011, emphasized enhanced item visibility, a reduction in perceived stress during task execution, and greater ease of accessing frequently used items as key benefits. These participants generally aligned with the “performers” or “motivated but physically strained” typologies observed in the quantitative profiles.

Conversely, participants who expressed negative or neutral views, such as ID0001, ID0002, ID0006, and ID0010, cited concerns regarding potential dust accumulation, disruption of kitchen aesthetics, loss of privacy of items stored in kitchen cabinets, and long-standing habits favoring closed cabinets. These participants often overlapped with the “struggler” or “motivated but physically strained” categories. Table 3 summarizes participants’ shelving preferences, key emergent themes from the interviews, and illustrative quotes that highlight their perspectives.

Five of the eleven participants expressed a favorable attitude toward open shelving design, whereas the remaining participants were either opposed to or ambivalent about adopting this design approach. These qualitative findings underscore the complexity of kitchen design interventions, where objective performance gains do not always align with subjective user acceptance.

**Table 6.** Participants’ Qualitative Perception.

| Participant ID | Desire for Open Shelving | Key Themes | Highlighted Quote |
| --- | --- | --- | --- |
| 0005 | Yes | Visibility, Stress Reduction | “I like my open pantry; it’s less stressful because I can see everything, especially with more items to remember.” |
| 0007 | Yes | Memory Aid, Visibility | “I can just turn around and see where everything is, instead of remembering which door it’s behind.” |
| 0009 | Yes | Ease of Access | “I think open shelving will be very helpful.” |
| 0011 | Yes | Ease of Access | “I have some open shelves, but most are closed, and I’d like more because it’s easy to get in and out.” |
| 0003 | Neutral | Safety, Privacy | “I’m not sure... Things might slide out of the cabinet, and I want my cabinets private, without dust or flies.” |
| 0004 | Neutral | Aesthetics | “I thought I’d like open shelving at first, but now I don’t think so because of the aesthetics.” |
| 0001 | No | Dust, Pet Concerns | “I’m afraid it’ll get dusty, especially since I have a dog.” |
| 0002 | No | Aesthetics, Privacy | "I like my kitchen tidy, and I don't want people seeing what's inside my cabinets." |
| 0006 | No | Aesthetics, Effort | "Open shelving spoils my kitchen's decor, and opening a cabinet isn't that much effort." |
| 0008 | No | Aesthetics | "I don't want open shelving." [No specific reason provided; implied aesthetic preference] |
| 0010 | No | Familiarity, Habit | "I've been in my house 30 years and already know where everything is." |

### Integration of Quantitative and Qualitative Findings

Analysis revealed that subjective preferences for open shelving did not always correspond to objective improvements in cognitive load, intrinsic motivation, or physical activity levels. For instance, participant ID0005 demonstrated both functional benefits and positive subjective experiences, supporting the potential effectiveness of open shelving for certain individuals. In contrast, participant ID0002 exhibited objective improvements in motivational and functional domains but remained resistant to open shelving, primarily due to aesthetic concerns.

These discrepancies highlight the importance of integrating functional, cognitive, and emotional dimensions when designing kitchen environments for older adults with MCI. Functional independence alone is insufficient; emotional attachment, aesthetic preferences, and habitual behaviors must be considered to foster true adoption and sustained use of environmental modifications.

## Discussions

This pilot study examined the impacts of open shelving versus closed cabinets on cognitive load, intrinsic motivation, physical effort, and user perceptions during kitchen tasks among older adults with MCI. The findings partially support the hypotheses derived from Cognitive Load Theory (CLT) and Self-Determination Theory (SDT), suggesting that visually accessible environments may yield subtle but meaningful benefits for cognitive processing, motivation, physical efficiency, and task duration.

Group-level analyses demonstrated improvements in several domains attributable to the open shelving condition. Specifically, participants exhibited decreased blink latency under open shelving, suggesting enhanced visual information processing and reduced cognitive effort in line with CLT principles (Sweller, 2010). Improvements in intrinsic motivation were reflected in increased perceived competence and reduced pressure/tension scores, aligning with SDT’s proposition that environmental supports promoting autonomy and competence foster greater intrinsic engagement (Ryan & Deci, 2000). Notably, physical activity data revealed a statistically significant reduction in step count with open shelving, indicating greater navigational efficiency when items were visually accessible. This latter finding is consistent with universal design principles that emphasize intuitive accessibility (Centre for Excellence in Universal Design, 2015).

The findings related to cognitive load, however, were mixed. While blink latency decreased, indicative of potentially lower cognitive strain, mean fixation duration slightly increased under the open shelving condition. This result may reflect a potential trade-off, where increased global visibility improved object retrieval but also concurrently introduced a degree of visual clutter, requiring longer fixations to identify relevant items. Such effects have been observed in prior studies examining MCI navigation in visually complex environments (Wang et al., 2022). The small sample size (n = 11) and inherent variability often observed in eye-tracking responses during experimental physiological monitoring (Holmqvist et al., 2011) likely contributed to the inconsistent cognitive load findings.

Despite measurable functional improvements, the subjective acceptance of open shelving was more nuanced and varied among participants. Qualitative data from post-task interviews revealed that while some participants valued the increased visibility and reduced cognitive demands associated with searching for items, others expressed resistance. This reluctance was often attributed to concerns about aesthetics, privacy preferences, and fears of environmental disorder. This divergence between objective benefits and subjective appraisal illustrate that cognitive-functional improvements alone may not drive adoption; emotional, aesthetic, and behavioral factors are crucial determinants of environmental acceptability, in line with Environmental Gerontology frameworks emphasizing the psychological meaning of home environments (Wahl & Oswald, 2016).

## Limitations and Future Work

The small sample size, due to the nature of the population, limited statistical power. Eye-tracking data for two participants were missing due to technical disruptions, a recognized challenge in physiological measurement research (Holmqvist et al., 2011). Although the within-subjects design mitigated individual variability, the use of a lab kitchen, despite randomization of condition order and including of breaks, may not fully capture the ecological validity of participants’ habitual home environments (Wang et al., 2022). Furthermore, while critical for isolating specific design effects, the controlled arrangement of ingredients and utensils in the cabinets cannot fully replicate the typical complexity, such as diversity and varying levels of clutter, found in real-world home kitchens. Finally, the limited racial, demographic, and cultural diversity of the sample constrains the generalizability of the findings to broader MCI populations.

Future research should expand sample sizes and diversify participant demographics to enhance the robustness and applicability of the findings. Studies conducted in participants’ homes would provide more ecologically valid insights into how visual accessibility affects everyday functioning. Investigating dynamic or adaptable shelving systems, such as polymer-dispersed liquid crystal (PDLC) technologies that toggle between transparency and opacity, may offer flexible solutions that balance accessibility with aesthetic and privacy concerns. Longitudinal studies are also needed to determine whether sustained exposure to open shelving or other visually accessible environments yields cumulative cognitive, motivational, and functional benefits over extended periods (Czaja et al., 2018).

## Conclusions

This study contributes preliminary evidence that enhancing visual accessibility in kitchen environments through open shelving can modestly reduce cognitive load, bolster intrinsic motivation, and improve physical navigation during kitchen tasks among older adults with MCI. The statistically significant decrease in step count and improvements in perceived competence highlight the potential for relatively simple design interventions to support functional independence meaningfully.

However, these findings also underscore the nuanced complexity in designing environments for cognitive aging. Emotional attachment to traditional aesthetics, long-standing habits, and subjective perceptions substantially influence the acceptance of environmental modifications, often overriding objective performance gains. Therefore, interventions aimed at supporting aging-in-place through kitchen design must not only prioritize cognitive and physical optimization but also sensitively address and honor the psychosocial dimensions of home environments.

Future research should pursue longitudinal, ecologically valid studies that incorporate flexible design solutions, such a PDLC, to better support aging individuals’ diverse needs. Bridging universal design principles with user-centered personalization remains critical to advancing living environments that both accommodate and empower older adults experiencing cognitive decline.

## Data Availability

All data produced in the present study are available upon reasonable request to the authors.

## Acknowledgements

We thank Jennifer DuBose, Zhi Tan, Bolaji Omofojoye, Alexander Adams, Katie Schreiber, Elahn Little, Madhuparna Sastakar, Abdurrahman Baru, and Marwan Shagar for their invaluable contributions to this study. We also thank the members, care partners, and staff at the Charlie and Harriet Shaffer Cognitive Empowerment Program and the Emory Brain Health Center for participating in this study and supporting our research. We also thank our colleagues and staff at the Georgia Institute of Technology, Emory University, and Northeastern University for their invaluable partnership.

## References

Anguita, D., Ghio, A., Oneto, L., Parra, X., & Reyes-Ortiz, J. L. (2013). A public domain dataset for human activity recognition using smartphones. Proceedings of the European Symposium on Artificial Neural Networks, 437–442.

Alzheimers.gov. (2024). What is mild cognitive impairment? https://www.alzheimers.gov/alzheimers-dementias/mild-cognitive-impairment

Binette, J. (2021, November). 2021 Home and Community Preferences Survey: A National Survey of Adults Age 18-Plus. Retrieved from https://www.aarp.org/research/topics/community/info-2021/2021-home-community-preferences.html.

Bland, J. M., & Altman, D. G. (1996). Transforming data. BMJ (Clinical research ed.), 312(7033), 770. 10.1136/bmj.312.7033.770

Cassarino, M., & Setti, A. (2016). Complexity as key to designing cognitive-friendly environments for older people. Frontiers in Psychology, 7, 1329. 10.3389/fpsyg.2016.01329

Centre for Excellence in Universal Design (2015). Universal Design Guidelines: Dementia Friendly Dwellings for People with Dementia, their Families and Carers. National Disability Authority. https://universaldesign.ie/Built-Environment/Housing.

Czaja, S. J., Boot, W. R., Charness, N., & Rogers, W. A. (2018). Designing for older adults: Principles and creative human factors approaches (3rd ed.). CRC Press.

DelPreto, J., Liu, C., Luo, Y., Foshey, M., Li, Y., Torralba, A., … & Rus, D. (2022). ActionSense: A multimodal dataset and recording framework for human activities using wearable sensors in a kitchen environment. Advances in Neural Information Processing Systems, 35, 13800–13813.

Derbie, A. Y., Dejenie, M. A., & Zegeye, T. G. (2022). Visuospatial representation in patients with mild cognitive impairment: Implication for rehabilitation. Medicine, 101(44), e31462. 10.1097/MD.0000000000031462

Efird, J. (2011). Blocked Randomization with Randomly Selected Block Sizes. International Journal of Environmental Research and Public Health, 8(1), 15–20. 10.3390/ijerph8010015

Ferraro, K., & Carr, D. (Eds.). (2021). Handbook of aging and the social sciences. Academic Press.

Freedson, P. S., Melanson, E., & Sirard, J. (1998). Calibration of the Computer Science and Applications, Inc. accelerometer. Medicine & Science in Sports & Exercise, 30(5), 777–781. 10.1097/00005768-199805000-00021

Grill, J. D., & Galvin, J. E. (2014). Facilitating Alzheimer disease research recruitment. Alzheimer disease and associated disorders, 28(1), 1–8. 10.1097/WAD.0000000000000016

Guralnik, J. M., & Ferrucci, L. (2003). Assessing the building blocks of function: utilizing measures of functional limitation. American journal of preventive medicine, 25(3), 112–121.

Guo, H. J., & Sapra, A. (2020). Instrumental activity of daily living.

Hollander, M., Wolfe, D. A., & Chicken, E. (2013). Nonparametric statistical methods (3rd ed.). Wiley.

Holmqvist, K., Nyström, M., Andersson, R., Dewhurst, R., Jarodzka, H., & Van de Weijer, J. (2011). Eye tracking: A comprehensive guide to methods and measures. Oxford University Press.

King, P. Y. A. (2023). Universal design in public housing: An ethnographic study on enhancing the quality of life of older people with mild cognitive impairment living alone.

Lakens, D. (2013). Calculating and reporting effect sizes to facilitate cumulative science: A practical primer for t-tests and ANOVAs. Frontiers in Psychology, 4, 863. 10.3389/fpsyg.2013.00863

Liao, Y.-Y., Chen, I.-H., Lin, Y.-J., Chen, Y.-H., & Tseng, H.-Y. (2021). Cognitive training enhances brain activation efficiency during working memory tasks in older adults. Nature Aging, 1(10), 869–879. 10.1038/s43587-021-00118-5

Liverman, C. T., Yaffe, K., & Blazer, D. G. (Eds.). (2015). Cognitive aging: Progress in understanding and opportunities for action.

Lussier, M., Adam, S., Chikhaoui, B., Consel, C., Gagnon, M., Gilbert, B., … & Bier, N. (2019). Smart home technology: a new approach for performance measurements of activities of daily living and prediction of mild cognitive impairment in older adults. Journal of Alzheimer’s Disease, 68(1), 85–96.

Machry, H., Motamed Rastegar, R., Gholami, Y., Yang, E., Little, E., Burke, M. A. M., … Zimring, C. (2025). Cognitive-Aging-in-Place: Home Design Factors Influencing Instrumental Activities of Daily Living for Older Adults Facing Mild Cognitive Impairment. Journal of Aging and Environment, 1–27. 10.1080/26892618.2025.2506060

McAuley, E., Duncan, T., & Tammen, V. V. (1989). Psychometric properties of the Intrinsic Motivation Inventory in a competitive sport setting: A confirmatory factor analysis. Research Quarterly for Exercise and Sport, 60(1), 48–58. 10.1080/02701367.1989.10607413

Mynatt, E. D., Essa, I., & Rogers, W. (2000). Increasing the opportunities for aging in place. In Proceedings of the 2000 Conference on Universal Usability (pp. 65-71). ACM. 10.1145/355460.355476

Morris, J. C., Storandt, M., Miller, J. P., McKeel, D. W., Price, J. L., Rubin, E. H., & Berg, L. (2001). Mild cognitive impairment represents early-stage Alzheimer disease. Archives of Neurology, 58(3), 397–405. 10.1001/archneur.58.3.397

Nagi, S. Z. (1976). An epidemiology of disability among adults in the United States. The Milbank Memorial Fund Quarterly. Health and Society, 439-467.

Nasreddine, Z. S., Phillips, N. A., B.dirian, V., Charbonneau, S., Whitehead, V., Collin, I., Cummings, J. L., & Chertkow, H. (2005). The montreal cognitive assessment, MoCA: A brief screening tool for mild cognitive impairment. Journal of the American Geriatrics Society, 53(4), 695–699. 10.1111/j.1532-5415.2005.53221.x

Petersen, R. C., Smith, G. E., Waring, S. C., Ivnik, R. J., Tangalos, E. G., & Kokmen, E. (1999). Mild cognitive impairment: Clinical characterization and outcome. Archives of Neurology, 56(3), 303–308. 10.1001/archneur.56.3.303

Rodrigues, F., Teixeira, D. S., Neiva, H. P., & Monteiro, D. (2023). The role of intrinsic motivation in promoting physical activity and well-being among older adults: A systematic review. Journal of Aging and Physical Activity, 31(2), 312–325. 10.1123/japa.2022-0156

Royston, P. (1995). Remark AS R94: A remark on Algorithm AS 181: The W-test for normality. Applied Statistics, 44(4), 547–551. 10.2307/2986146

Ryan, R. M. (1982). Control and information in the intrapersonal sphere: An extension of cognitive evaluation theory. Journal of Personality and Social Psychology, 43(3), 450–461. 10.1037/0022-3514.43.3.450

Ryan, R. M., & Deci, E. L. (2000). Self-determination theory and the facilitation of intrinsic motivation, social development, and well-being. American Psychologist, 55(1), 68–78. 10.1037/0003-066X.55.1.68

Salvucci, D. D., & Goldberg, J. H. (2000). Identifying fixations and saccades in eye-tracking protocols. Proceedings of the 2000 Symposium on Eye Tracking Research & Applications (ETRA ’00), 71–78. 10.1145/355017.355028

Stephens, M. A. (1974). EDF statistics for goodness of fit and some comparisons. Journal of the American Statistical Association, 69(347), 730–737. 10.2307/2286009

Suresh K. (2011). An overview of randomization techniques: An unbiased assessment of outcome in clinical research. Journal of human reproductive sciences, 4(1), 8–11. 10.4103/0974-1208.82352 (Retraction published J Hum Reprod Sci. 2023 Jan-Mar;16(1):87)

Sullivan, G. M., & Feinn, R. (2012). Using effect size—or why the p-value is not enough. Journal of Graduate Medical Education, 4(3), 279–282. 10.4300/JGME-D-12-00156.1

Sweller, J. (2010). Cognitive load theory: Recent theoretical advances. In J. L. Plass, R. Moreno, & R. Brünken (Eds.), Cognitive load theory (pp. 29–47). Cambridge University Press.

Tan, J. P., Li, N., Gao, J., Wang, L. N., Zhao, Y. M., Yu, B. C., Du, W., Zhang, W. J., Cui, L. Q., Wang, Q. S., Li, J. J., Yang, J. S., Yu, J. M., Xia, X. N., & Zhou, P. Y. (2015). Optimal cutoff scores for dementia and mild cognitive impairment of the Montreal Cognitive Assessment among elderly and oldest-old Chinese population. Journal of Alzheimer’s disease : JAD, 43(4), 1403–1412. 10.3233/JAD-141278

Thomas, W., & Blanchard, J. (2009). Moving beyond place: Aging in community. Generations, 33(2), 12–17.

Vickers, K., Rodriguez, A., Goldstein, F., Ariyo, A., Binford, S., Campiglia, G., … & Levey, A. (2020). Cognitive Empowerment Program for Individuals with Mild Cognitive Impairment: Description and Preliminary Satisfaction Ratings. Archives of Physical Medicine and Rehabilitation, 101(11), e107.

Wang, L., Lin, C.-C., & Huang, Y.-H. (2022). Cognitive and physical challenges in the kitchen: Implications for older adults with mild cognitive impairment. Aging & Mental Health, 26(8), 1567–1574. 10.1080/13607863.2021.1922601

Willis, S. L., Tennstedt, S. L., Marsiske, M., Ball, K., Elias, J., Koepke, K. M., … & Wright, E. (2009). Long-term effects of cognitive training on everyday functional outcomes in older adults. Journal of the American Medical Association, 296(23), 2805–2814. 10.1001/jama.296.23.2805

Zagermann, J., Pfeil, U., & Reiterer, H. (2016). Measuring cognitive load using eye-tracking technology in visual computing. Proceedings of the Sixth Workshop on Beyond Time and Errors on Novel Evaluation Methods for Visualization, 78–85. 10.1145/2993901.2993908

